# Perisynaptic matrix gene variants underlie DystoniaGeneNet endophenotypes in isolated focal dystonia

**DOI:** 10.64898/2026.10.19.26365640

**Authors:** Aloysius Domingo, Dongren Yao, Nyah Kshatriya, Kotchaphorn Mangkalaphiban, Giovanni Battistella, Lena C. O’Flynn, Bailey Reese, Madison Hincher, Rachita Yadav, Nutan Sharma, Laura Williams, Conor Fearon, Owen Killian, Richard Reilly, Michael Hutchinson, Deborah Raymond, Rachel Saunders-Pullman, Susan Bressman, Steven J. Frucht, Michael Talkowski, Kristina Simonyan, Laurie J. Ozelius

## Abstract

Isolated focal dystonia is a rare movement disorder characterized by abnormalities in brain structure and function. The genes underlying isolated focal dystonia are not fully elucidated, and genetic studies are constrained by incomplete penetrance and small family sizes. Neuroimaging endophenotypes shared by patients and unaffected relatives can reveal genetic liability independently of clinical manifestation and offer a strategy for advancing gene discovery. We developed DystoniaGeneNet, a 3D convolutional neural network that predicts familial status from T1-weighted brain MRIs. We then used its predictions to guide genetic analyses in isolated focal dystonia. DystoniaGeneNet was trained on 40 patients with familial laryngeal dystonia and 40 healthy controls, then applied to independent cohorts of 21 patients with familial isolated focal dystonia, 364 patients with sporadic dystonia, and 98 unaffected relatives. Model predictions were incorporated into variant prioritization and segregation analysis in 214 individuals with whole-exome sequencing data, and results were replicated in another 738 independent exomes, in genetic data from a hospital-based biobank (892 dystonia cases, 2,475 controls), and in the *All of Us* Research Program genetic data (1,343 dystonia cases, 6,840 controls). DystoniaGeneNet discriminated familial dystonia from healthy controls in the training set and correctly classified 85.7% of independent familial cases, using a biomarker centred on the corpus callosum and inferior temporal gyrus. Across the discovery and replication cohorts, we identified rare, likely pathogenic genetic variants in *RAP1GAP* and *TNN* segregating with disease in families. Additional variants were found in singletons with isolated focal dystonia. In patients with clinically sporadic dystonia but predicted to have familial features, we observed multiple rare variants in *ITSN2*, including three high-confidence loss-of-function variants, and an enrichment of damaging missense variants in its Dbl homology domain. To determine the functional impact of discovered genetic variants, we assessed gene expression and splicing via qPCR and RNA sequencing (*n* = 74), revealing reduced *RAP1GAP* and *ITSN2* expression in variant carriers. At the pathway level, rare nonsynonymous variants in brain-expressed matrisome genes were associated with dystonia (*P* = 0.012). Our strategy utilizing a deep learning-derived neuroimaging endophenotype to empower gene discovery uncovered *RAP1GAP, TNN*, and *ITSN2* as novel genes associated with isolated focal dystonia. These genes converge on perisynaptic extracellular matrix-synapse signalling, implicating a mechanism linked to synaptic plasticity deficits, and identifying a molecular substrate for network-level brain abnormalities that characterize this disorder. Further, we exemplify a generalizable framework for leveraging endophenotypes in rare neurogenetic diseases with incomplete penetrance.

## Introduction

Isolated focal dystonia is a rare neurological movement disorder characterized by abnormal muscle tone and sustained or intermittent muscle contractions, resulting in involuntary spasms and/or twisting postures.^1^ It affects about 30 per 100,000 individuals in the population,^2^ and significantly impairs quality of life for patients.^3^ Current treatments are predominantly limited to symptom management; however, recent studies have begun to clarify the pathophysiologic underpinnings of dystonia, informing paths towards mechanistic therapeutic strategies.^4^

The genetic architecture of dystonia encompasses disruption of genes causative for isolated forms as well as genes implicated in syndromic conditions. Approximately 50 genes associated with monogenic dystonia have been identified,^5^ but only a small fraction (fewer than five genes) are associated with isolated focal dystonia.^6,7^ The likelihood of identifying a genetic etiology for dystonia is highest in individuals with early age-at-onset and family history.^8,9^ Nevertheless, a substantial proportion of familial cases remain unexplained, highlighting opportunities for novel gene discovery.

Several factors have thus far constrained genetic studies in dystonia. It is a rare disorder with marked phenotypic variability and a broad age-at-onset distribution, complicating the collection of large, clinically homogeneous cohorts that are ideal for association studies.^10^ Pleiotropy, in which a gene is associated with multiple seemingly unrelated phenotypes, poses challenges for variant interpretation.^9^ Finally, incomplete penetrance is an almost invariant feature across inherited forms of dystonia.^11^ As carriers of pathogenic variation in dystonia-associated genes do not always manifest disease, this often results in sparsely affected pedigrees and ineffective segregation studies.

One strategy to overcome these challenges is to leverage an endophenotype, a measurable biomarker that effectively discriminates at-risk from truly unaffected individuals based on genetic liability instead of outward disease manifestation.^12^ Because an endophenotype is more penetrant than the disease phenotype, it can be used to identify non-manifesting carriers (NMCs) in families, thus expanding the number of individuals available for segregation analyses. Moreover, in sporadic disease, where a subset of singletons plausibly carries monogenic disease variants but with strikingly reduced penetrance,^13,14^ identifying genetic liability through biomarkers may offer an alternative route to gene discovery beyond family-based approaches.

Prior neuroimaging studies have shown that abnormalities in sensorimotor brain regions relevant to dystonia pathophysiology are shared between symptomatic patients and their unaffected relatives,^15^ indicating that these can be used as endophenotypes. Further, neural features derived from brain images can discriminate between inherited and sporadic dystonia,^16^ as well as between dystonia patients and NMCs versus non-susceptible healthy controls,^15^ suggesting that these are heritable traits and proxy subclinical genetic etiology and pathogenesis. In large association studies for complex traits, deep learning-based neuroimaging endophenotypes captured the genetic architecture of heritable brain structural changes.^17^

In this study, we hypothesized that neuroimaging endophenotypes could be exploited in a rare disease to systematically aid gene discovery. To this end, we developed a deep learning model, DystoniaGeneNet, which was trained on structural brain images to predict the probability of familial dystonia. We reasoned that, in a relatively homogeneous cohort of adult-onset isolated focal dystonia, individuals with a high probability of familial status would have the strongest likelihood of a genetic etiology. As such, we used DystoniaGeneNet to label predicted NMCs (pNMCs) in families and refine the number of informative individuals for segregation analysis. In singletons, we used the model to predict which individuals were likely carriers of causative genetic variation. Using this approach, we identified three new causative genes for isolated focal dystonia. Moreover, we uncover an association between dystonia and the disruption of genes related to the perisynaptic extracellular matrix (ECM), a pathway that is relevant to dystonia pathogenesis with its link to synaptic plasticity deficits and disordered brain network connectivity.^18,19^

## Materials and Methods

### Study design

#### Cohorts

To determine the genetic underpinnings of isolated focal dystonia through neuroimaging endophenotypes, we assembled several cohorts and datasets (**Figure 1**):

**Figure 1.**
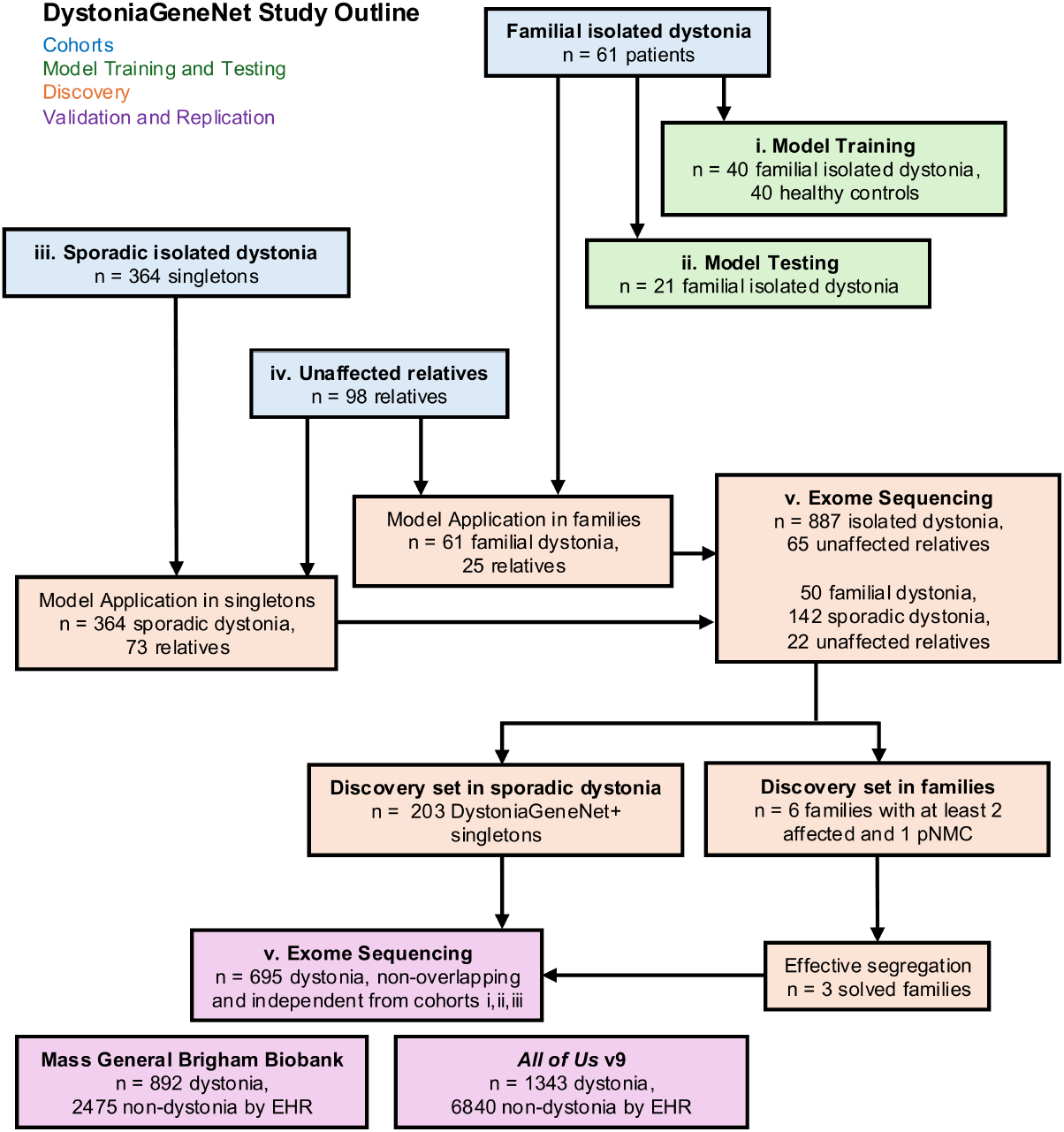
Study Outline. Clinically sporadic individuals (singletons) were followed-up longitudinally for development of dystonia in a family member (sporadic to familial phenoconversion).

i. the DystoniaGeneNet training cohort of 40 patients with familial laryngeal dystonia (LD) and 40 healthy controls;
ii. an independent testing cohort of 21 patients with familial dystonia;
iii. an independent testing cohort of 364 patients with sporadic LD and other forms of isolated focal dystonia;
iv. an independent cohort of 98 unaffected relatives of dystonia patients; and
v. a dataset of 952 whole-exome sequences (WES), including 887 patients with isolated dystonia and 65 unaffected relatives;

Clinical and demographic features are found in **Table 1**. Patients with clinically sporadic dystonia and unaffected relatives were longitudinally monitored through yearly telephone interviews to assess whether there was development of dystonia within their families (sporadic to familial phenoconversion).

**Table 1.** Cohort demographics.

| *Cohort i – Model Training* |  |  |
| --- | --- | --- |
|  | **40 patients with isolated familial dystonia** | **40 healthy volunteers** |
| **Phenotypes** | 39 LD, 1 multifocal dystonia | n/a |
| **Age at scan** (years; mean ± std) | 58.15 ± 12.81 | 48.75 ± 11.07 |
| **Sex** (F/M) | 30 / 10 | 19 / 21 |
| **Disease duration** (years; mean ± std) | 19.73 ± 12.67 (n=39) | n/a |
| **Age of onset** (years; mean ± std) | 38.08 ± 15.70 (n=39) | n/a |
| **Imaging site** | 4 MGB, 1 NIH, 35 ISMMS | 9 NIH, 31 ISMMS |
| **MRI scanner** (3T, MPAGE) | 1 GE, 20 Philips, 19 Siemens | 9 GE, 12 Philips, 19 Siemens |
| *Cohort ii – Independent Testing* |  |  |
|  | **21 patients with isolated familial dystonia** |  |
| **Phenotypes** | 17 LD, 2 WC, 1 BLS, 1 multifocal dystonia |  |
| **Age at scan** (years; mean ± std) | 59.67 ± 13.50 | |
| **Disease duration** (years; mean ± std) | 19.00 ± 14.74 | |
| **Age of onset** (years; mean ± std) | 40.67 ± 15.71 | |
| **Sex** (F/M) | 18 / 3 |  |
| **Imaging site** | 2 MGB, 18 ISMMS, 1 NIH |  |
| **MRI scanner** (3T, MPAGE) | 12 Philips, 8 Siemens, 1 GE |  |
| *Cohort iii – Sporadic/Singletons* |  |  |
|  | **364 patients with isolated sporadic dystonia** |  |
| **Phenotypes** | 291 LD, 19 WC, 15 BLS, 14 MFHD, 12 CD, 5 segmental dystonia, 4 Meige’s syndrome, 3 OMD, 1 multifocal dystonia |  |
| **Age** (years; mean ± std) | 55.35 ± 12.57 (n=363) | |
| **Sex** (F/M) | 256 / 108 |  |
| **Imaging site** | 142 MGB, 178 ISMMS, 44 NIH |  |
| **MRI scanner** (3T, MPAGE/MP2RAGE) | 104 Philips, 216 Siemens, 44 GE |  |
| *Cohort iv – Unaffected Relatives* |  |  |
|  | **98 unaffected relatives of isolated dystonia patients** |  |
| **Phenotypes of proband** | 94 LD, 2 segmental dystonia, 1 WC, 1 BLS |  |
| **Age** (years; mean ± std) | 46.45 ± 16.02 | |
| **Sex** (F/M) | 68 / 30 |  |
| **Imaging site** | 75 MGB, 23 ISMMS |  |
| **MRI scanner** (3T, MPAGE/MP2RAGE) | 91 Siemens, 7 Philips |  |
| *Cohort v – Whole-Exome Sequencing Dataset* |  |  |
|  | **887 patients with isolated dystonia** | **65 unaffected relatives** |
| **Dystonia distribution** | 843 focal or segmental<br>44 multifocal or generalized | n/a |
| **Sex** (F/M) | 651 / 236 | 41 / 24 |
| **Inheritance Pattern** | 352 familial<br>527 sporadic, 8 unknown | n/a |
| **Genetic Ancestry Groups** | 891 European, 30 African, 16 American, 15 other ancestries |  |

BLS – Blepharospasm; CD – Cervical dystonia; F – female; ISMMS – Icahn School of Medicine at Mount Sinai; LD – Laryngeal dystonia; M – male; MFHD – Musician’s focal hand dystonia; MGB – Mass General Brigham; MPAGE – Magnetization prepared rapid gradient echo; MP2RAGE – Magnetization prepared two rapid acquisition gradient echo; NIH – National Institute of Health; OMD – Oromandibular dystonia; WC – Writer’s cramp

#### Inclusion and exclusion criteria

Diagnoses and phenotypes of dystonia, as well as the absence of other neurological and psychiatric diagnoses, were established based on detailed case history, review of medical information, and neurological or laryngeal evaluations. None of the participants received any centrally acting medications or had undergone surgery of the brain, head, or neck at the time of participation for the neuroimaging component of the study. Family history of dystonia was established by the presence of at least one blood relative with a confirmed diagnosis of dystonia. Patients harbouring pathogenic variants in genes associated with isolated dystonia (*TOR1A* GAG deletion, likely pathogenic missense variants in *THAP1, GNAL, ANO3, KMT2B*, and loss-of-function variants in *VPS16)* were excluded whenever genetic data were available. This study was approved by the Institutional Review Board (IRB) of Mass General Brigham. Cohort collection for genetic studies was approved by IRBs of Icahn School of Medicine at Mount Sinai and University College Dublin. All participants gave written informed consent prior to study participation in accordance with the Declaration of Helsinki.

### Model development and training

Whole-brain T1-weighted images were acquired on a 3.0 Tesla MRI scanner using 3D magnetization-prepared rapid gradient echo (MPRAGE) or magnetization-prepared 2 rapid acquisition gradient echo (MP2RAGE) sequence protocols. Training and testing sets were balanced to minimize potential effects of scanner hardware, data collection sites, and head coils (**Table 1**). Head movements during scanning were minimized using padding between the coil and the participant’s head. Images were manually inspected for their quality to ensure the absence of artifacts. We applied a streamlined, semi-automated preprocessing pipeline before training DystoniaGeneNet. Each scan was first reoriented to the anterior commissure–posterior commissure (AC-PC) line, followed by skull stripping using a U-Net based approach, N4 bias field correction, and affine registration to the MNI152 template. The registered images were then resampled to an isotropic resolution of 1 × 1 × 1 mm^3^ with a final matrix size of 181 × 217 × 181 mm^3^.

DystoniaGeneNet was built as a modified version of DystoniaNet^20^ using a 3D convolutional neural network (3D-CNN) architecture, comprising a feature-extraction module and a decision-making module, using raw structural brain MRIs. The feature extractor was based on a 3D ResNet-50 architecture with four residual stages (Res3D-1 to Res3D-4), containing 3, 4, 6, and 3 bottleneck blocks, respectively. All blocks shared the same internal layout of a convolution layer, batch normalization, and ReLU activation. Each block consisted of a 1 × 1 × 1 convolution for channel reduction, a 3 × 3 × 3 convolution for spatial feature extraction, and a final 1 × 1 × 1 convolution for channel expansion. The number of channels increased progressively across stages, following the standard ResNet-50 configuration: (64, 64, 256), (128, 128, 512), (256, 256, 1024), and (512, 512, 2048). Residual shortcut pathways were incorporated to facilitate optimization and preserve feature propagation across depth; where dimensional mismatch occurred, particularly at stage transitions, projection shortcuts were used instead of direct identity connections.

Following the convolutional layers, the resulting feature maps were compressed by adaptive 3D global max pooling and flattened into a compact representation. The classification module then applied a fully connected layer with 1D batch normalization, ReLU activation, and dropout (rate = 0.6), followed by a final fully connected layer with a SoftMax activation to estimate the probability of isolated familial dystonia for each subject.

DystoniaGeneNet was trained using the Adam optimizer for 150 epochs with a batch size of 4. Early stopping was applied based on training loss over the final 40 epochs to prevent overfitting and improve efficiency. Classification was refined using a dynamic decision range defined by two thresholds (t_*f*_ and t_*nf*_), as described previously.^20^ The model produced one of three possible predictive outcomes for familial dystonia status: Yes, No, or Referral. A case with a probability of t_*f*_ > 0.52 was classified as a carrier of familial dystonia status, whereas a case with a probability of t_*nf*_ < 0.42 was classified as non-familial. Cases with a probability of t_*r*_ = 0.42-0.52 were referred, constrained to no more than 10% of all predictions. All training and testing were performed on an NVIDIA Tesla A6000 GPU with 48 GB of memory.

### Genomic and bioinformatic methods

#### Whole-exome sequencing, quality control, and preprocessing

Whole-exome sequencing (WES) libraries were constructed using Agilent SureSelect Human All Exon kits. Raw sequences were generated using paired-end sequencing (Illumina). WES preprocessing was performed using GATK workflows (Genome Analysis Toolkit, Broad Institute) in Terra.^21^ Briefly, raw FASTQ files were converted to unmapped BAMs (uBAM), cleaned, then mapped to the GRCh38 human genome reference (versions used: GATK version 4.2.6.1, Picard version 2.26.10). Samples were then jointly called and genotyped (Exome and JointGenotyping workflows versions v3.0.0 and v1.6.8), resulting in a cohort-level variant call file (VCF). Each VCF was then hard-filtered using custom GATK scripts, with the following parameters: for biallelic singlenucleotide variants (SNVs), variants were filtered out when QD < 3.0, MQ < 50.0, SOR > 2.5, ReadPosRankSum < -1.4, DP < 10 || DP > 1000; for indels, variants were filtered out when QD < 4.0, MQ < 50.0, SOR > 3.0, ReadPosRankSum < -1.7, DP < 10 || DP > 1000.

Variant calls were filtered to obtain family-wise files, and then segregating variants were determined using GATK modules. Annotation was performed using Ensembl Variant Effect Predictor v115 (www.ensembl.org/vep) and the database for nonsynonymous SNPs’ functional predictions (dbNSFP version 5.3). Population allele frequencies (AF) were drawn from the Genome Aggregation Database (gnomAD v4.1-joint) and from genetic data from the *All of Us* Research Program (v9).

#### Genetic ancestry inference

Genetic ancestry in WES samples was inferred using the gnomAD v4.0 PCA projection and pretrained random forest classifier. Exome variants were intersected with the gnomAD v4.0 PCA loading variants, yielding 81,156 variants for projection. Genotypes were quality-filtered using the default gnomAD adj criteria. Samples were projected onto the gnomAD PCA space using the publicly available PCA loadings and allele frequencies, and the first 20 projected PCs were used for ancestry classification. For broad ancestry assignment, probabilities for NFE, FIN, ASJ, and AMI were summed to obtain a European (EUR) probability. Samples were assigned to the broad ancestry group with the highest probability among EUR, AFR, AMR, and other genetic ancestries (EAS, MID, and SAS). The workflow was obtained from https://gnomad.broadinstitute.org/news/2021-09-using-the-gnomad-ancestry-principal-com ponents-analysis-loadings-and-random-forest-classifier-o n-your-dataset/.

#### Variant filtration and prioritization

After incorporating case-by-case DystoniaGeneNet probabilistic classification in segregation analysis, variants were prioritized into “likely pathogenic”, “likely benign” or “ambiguous” categories using a decision framework combining predicted impact, constraint, and population AF. Specifically, loss-of-function (LoF) variants (stop-gain, frameshift, splice donor, splice acceptor) were classified by LOFTEE^22^ as high-confidence or low-confidence, while missense variants received a pathogenicity score (MisPath) composed of CADD (Combined Annotation-Dependent Depletion),^23^ AlphaMissense^24^ and REVEL (Rare Exome Variant Ensemble Learner)^25^ tiers, binned into integer scores from −4 to +4 based on recommended calibrated thresholds.^26^ Gene-level constraint was assessed using gnomAD LOEUF (LoF observed/expected upper-bound fraction).^22^

High priority likely pathogenic variants included ultrarare variants (gnomAD and *All of Us* population AF ≤ 2×10^-5^) that were high-confidence LoF variants in highly constrained genes (LOEUF ≤ 0.45), missense variants with MisPath ≥ 1 in a constrained gene or region, or missense variants with MisPath ≥ 4 irrespective of constraint. Secondary priority was assigned to ultrarare variants that were either high-confidence LoF in moderately constrained genes (0.45 < LOEUF ≤ 1) or missense variants with MisPath ≥ 1 outside constrained regions. Variants were classified as likely benign if they were low-confidence LoF, LoF in unconstrained genes (LOEUF > 1), missense with MisPath < 0, or 5’/3’ UTR variants without elevated CADD (CADD-Phred score < 25). Any variant with population AF that is more common (gnomAD or *All of Us* AF > 5.0×10^-5^) was also classified as likely benign regardless of other features. Variants that exceeded the ultrarare threshold but remained below the more common population frequency threshold were also classified as likely benign, but were moved to ambiguous if they were (i) missense variants with MisPath ≥ 4, or (ii) high-confidence LoF variants in constrained genes (LOEUF ≤ 1), on the rationale that their population frequency is too elevated to support a pathogenic prioritization but their predicted impact is too strong to confidently exclude. All remaining variants, in-frame indels, missense with MisPath = 0, and borderline cases not meeting any tier criteria were classified as ambiguous for manual review.

### Replication in biobanks

Additional genetic evidence was obtained from two biobanks with genomic sequencing data: the in-house hospital-based Mass General Brigham Biobank (MGBB), a biorepository of consented patient samples at Mass General Brigham (Boston, MA, USA), and the US-based population biobank *All of Us* Research Program dataset.^27^

In the MGBB, individuals with dystonia were extracted using electronic health records (EHR)-based International Classification of Diseases (ICD-10) codes for dystonia (G24.1-9), excluding drug-induced dystonia (G24.0), and were restricted to individuals of self-reported White race and non-Hispanic ethnicity (587 females, 305 males). Manual case review was performed for individuals identified to harbour genetic variants in *RAP1GAP, TNN*, and *ITSN2*. Additionally, a set of White and non-Hispanic individuals with no evidence of neurologic disease (exclusion of G00-G99) was used as controls (1,313 females, 1,162 males).

We also used the *All of Us* Research Program Controlled Tier v9 Curated Data Repository (CDR), available through the Researcher Workbench. Phenotypes were derived from the Observational Medical Outcomes Partnership (OMOP) Common Data Model. Analyses were restricted to European participants with at least two distinct records of dystonia but no drug-induced or acquired dystonia (941 females, 402 males). A set of controls (3,881 females, 2,959 males) had no evidence of disorders of the nervous system, and were required to be at least 50 years old, with the reasoning that younger participants may not have yet manifested but are susceptible to adult-onset dystonia.

Genetic variation was queried using Hail version 0.2.135 on the *All of Us* Researcher Workbench. Analyses were performed on the Hail VariantDataset (VDS) generated from the CDR v9 joint-called SNP and indel WGS data aligned to GRCh38. For each gene, the VDS was filtered to the exonic intervals of the gene’s MANE Select transcript, with 10 base pairs of padding added at each exon boundary. The filtered VDS was converted to a dense Hail MatrixTable for genotype-level analysis. Only variants passing variant-level filters were retained, and genotype entries flagged as failed by entry-level filtering were masked. For each retained variant, alternate allele count, allele number, alternate allele frequency, and homozygous alternate genotype count were calculated in the full *All of Us* CDR v9 WGS cohort across all genetic ancestry groups. These overall allele frequency estimates were used for rare-variant filtering. Variants were annotated using Ensembl Variant Effect Predictor (VEP) v115 as with the dystonia exome cohort.

### Expression analyses

#### RNA sequencing and outlier analysis

RNA was extracted from blood collected from 74 individuals with dystonia (*n* = 54) and healthy controls (*n* = 20). From this, RNA-sequencing profiles were generated using the Illumina TruSeq Stranded library preparation kit with poly(A) enrichment and 150-bp paired-end sequencing. Raw sequencing reads were aligned to the reference genome (GRCh38) using the STAR v2.5.3 aligner. Visualization of BAM files was performed using Integrative Genomics Viewer v2.19.7.

To identify genes with aberrant expression, we performed outlier detection analysis on RNA sequencing count data us-ing OUTRIDER (Outlier in RNA-seq fInDER, v1.28.1).^28^ Lowly expressed genes were first removed by retaining only genes with a mean raw count greater than 10 and with detectable expression (count > 0) in at least 25% of samples, yielding 13,674 genes for analysis. The optimal autoencoder encoding dimension was estimated using the Optimal Hard Threshold (OHT) method via estimateBestQ(), which identified *q* = 13. The OUTRIDER model was then fitted with this *q* to correct for confounding factors and co-expression structure. Gene-level *P*-values were computed under a negative binomial distribution. Outlier calls for genome-wide discovery were made at a threshold of *P* < 0.05 and |Z-score| > 2. Analyses were performed in R 4.5.3.

#### Quantitative PCR

RNA was extracted from blood from a proband and three controls. Relative expression of *RAP1GAP* transcripts was measured using three TaqMan assays (Hs00937968_m1, Hs00939528_m1, Hs00939529_m1) and housekeeping genes (*GUSB, UBC, HPRT1*). Control RNA was obtained from unaffected individuals. 2^-ΔCT^ method was used to quantify relative expression. All experiments were performed in triplicate using standard two-step protocols on a QuantStudio 7 Pro machine.

### Protein domain enrichment

We used the *All of Us* case-control dataset for within-domain enrichment analyses. For each UniProt-defined protein domain of Rap1GAP, TNN, and ITSN2, missense variation with AF ≤ 0.01 were classified into rare functional (AlphaMissense score ≥ 0.17 and AF ≤ 1 × 10^-4^) versus other missense variants, then aggregate counts in dystonia and controls were determined. Contiguous segments were collapsed. For each domain, a 2×2 contingency table was constructed. Fisher’s exact test was applied, and *P*-values were adjusted using the Benjamini-Hochberg method.

### Pathway rare variant association analyses

Rare-variant association analysis was performed in the MGBB case-control dataset using Optimized Sequence Kernel Association Tests (SKAT-O) implemented in GMMAT (v1.4.2).^29^ A null logistic mixed model was fitted with dystonia as the outcome, the first five ancestry principal components as fixed-effect covariates, and a genomic relationship matrix as a random effect. Analyses were restricted to coding nonsynonymous variants within the matrisome gene set, refined to brain-expressed genes (*n* = 82 genes with transcripts per million ≥ 1 in at least one brain region, Genotype-Tissue Expression v8 dataset).^30^ The analysis excluded *RAP1GAP, TNN*, and *ITSN2* to maintain independence from previous discoveries. An aggregate statistic was also computed across all variants within the gene set and then compared in a competitive permutation test against 10,000 random gene sets of identical size, drawn without replacement from a background pool of genes that excluded matrisome genes.

## Results

### DystoniaGeneNet discovers neural features of familial dystonia

DystoniaGeneNet automatically recognized heritable anatomical features of familial isolated focal dystonia, classified individuals with a familial history, and predicted familial status in clinically sporadic cases (singletons) (**Figure 2A-D**).

**Figure 2.**
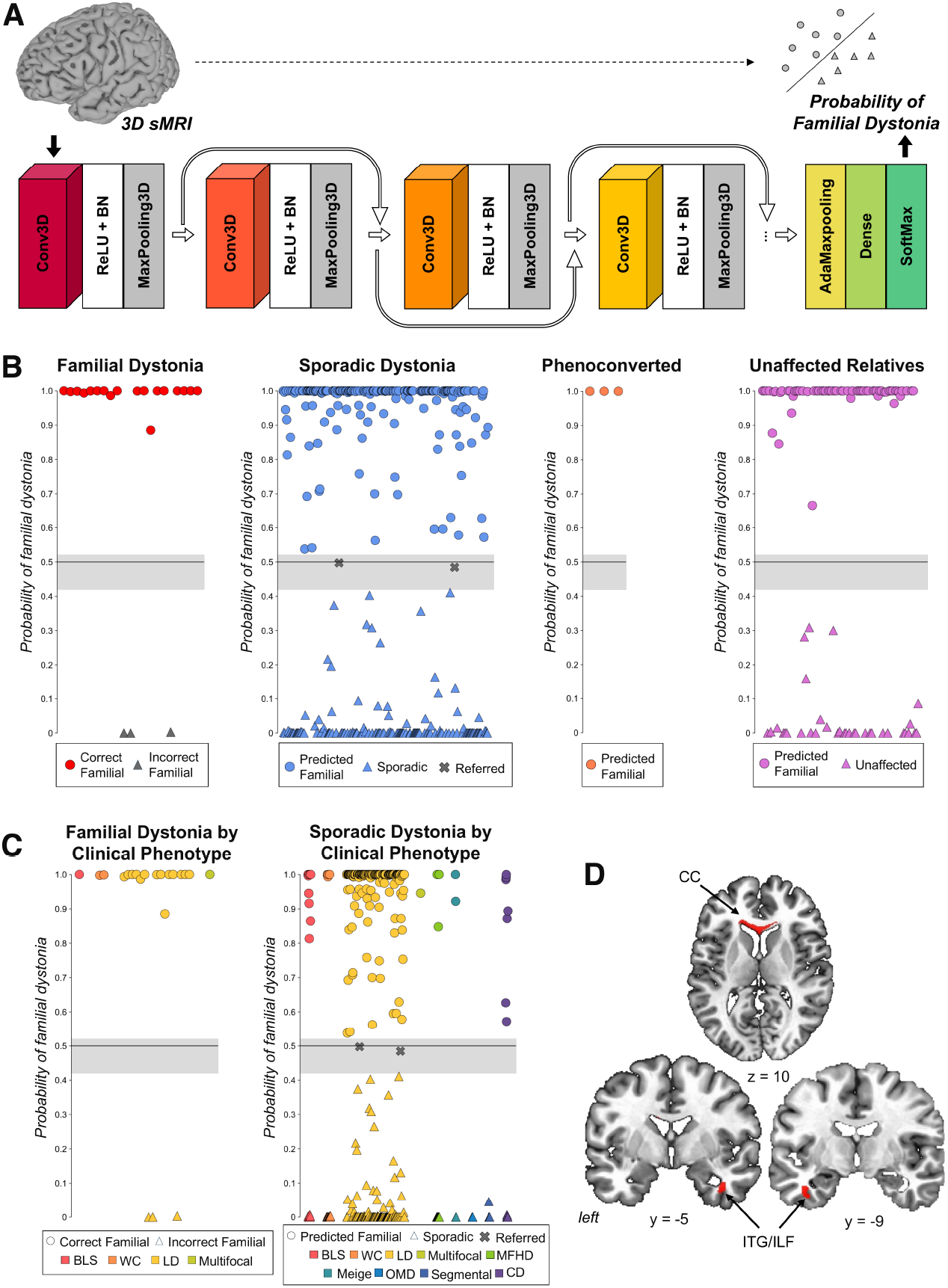
DystoniaGeneNet model architecture, performance, and biomarker visualization. **A**. Architecture of DystoniaGeneNet, a 3D convolutional neural network model with feature extraction and decision-making modules. Raw 3D structural brain magnetic resonance imaging (sMRI) data are model inputs, and individual probabilities of familial dystonia are the final model outputs. **B**. Overall performance of DystoniaGeneNet in an independent test set of familial dystonia patients, and distribution of predictions in independent cohorts of sporadic dystonia patients, phenoconverted cases, and unaffected relatives of dystonia patients. **C**. DystoniaGeneNet predictions in familial and sporadic dystonia patients by clinical phenotype. **D**. 2D visualization of the neural network biomarkers of DystoniaGeneNet defined as the top 15% of the voxelwise absolute maximum weight (in red) identified by the first convolutional block depicted on a series of axial and coronal brain slices in the Talairach-Tournoux standard space. BLS – blepharospasm, CC – corpus callosum, CD – cervical dystonia, ILF – inferior longitudinal fasciculus, ITG – inferior temporal gyrus, LD – laryngeal dystonia, MFHD – musician’s focal hand dystonia, OMD – oromandibular dystonia, sMRI – structural magnetic resonance imaging, WC – writer’s cramp.

The training model achieved 100% performance in discriminating patients with familial isolated focal dystonia from healthy individuals using a neural biomarker comprising the genu of the corpus callosum (CC) and the bilateral inferior temporal gyrus (ITG), extending to the inferior longitudinal fasciculus (ILF) (**Figure 2D**). In the independent testing set of patients with familial isolated focal dystonia, DystoniaGeneNet achieved a true predictive rate of 85.7%, correctly identifying 18 out of 21 familial cases. In the cohort of singletons, DystoniaGeneNet predicted 203 (55.8%) cases to be familial and 159 (43.7%) as truly sporadic, while referring 2 (0.5%) cases (**Figure 2B**). There were no evident trends in the model’s classification of different focal dystonia presentations across familial or sporadic cohorts (**Figure 2C**), indicating that while DystoniaGeneNet was trained primarily on laryngeal dystonia (LD), it could capture neural features in other presentations of isolated focal dystonia. Among unaffected relatives of patients with dystonia, 64 (65.3%) were classified as predicted NMCs (pNMC) and 34 (34.7%) as non-susceptible.

Longitudinal monitoring identified three cases of sporadic to familial phenoconversion, i.e., development of dystonia in a family member, prompting a change in the proband’s clinical classification. The average time between disease onset and phenoconversion was 20.7 ± 11.1 years. All three probands were accurately classified by DystoniaGeneNet as familial dystonia (average score of 1.0 ± 0.0), indicating that the model can be used to prospectively identify singletons with a high probability of carrying monogenic dystonia genes.

### Genetic variants in *RAP1GAP* and *TNN* identified in isolated dystonia families

We then leveraged DystoniaGeneNet classifications to refine the selection of individuals for segregation analysis in families. We selected six families to test this approach, based on the availability of two individuals affected with isolated focal dystonia in each family, and an additional family member confidently labelled (>0.90 probability) as a pNMC by the model (**Figure 3**). In three families, this approach identified a single likely pathogenic variant segregating with both dystonia and the endophenotype, and nominating a distinct causative gene in each family: *RAP1GAP, TNN*, and *TMEM190*. In three other families, variant prioritization did not lead to a putative gene; however, segregation analysis with DystoniaGeneNet classification effectively reduced the number of candidate genes (**Supplementary Table 1**).

**Figure 3.**
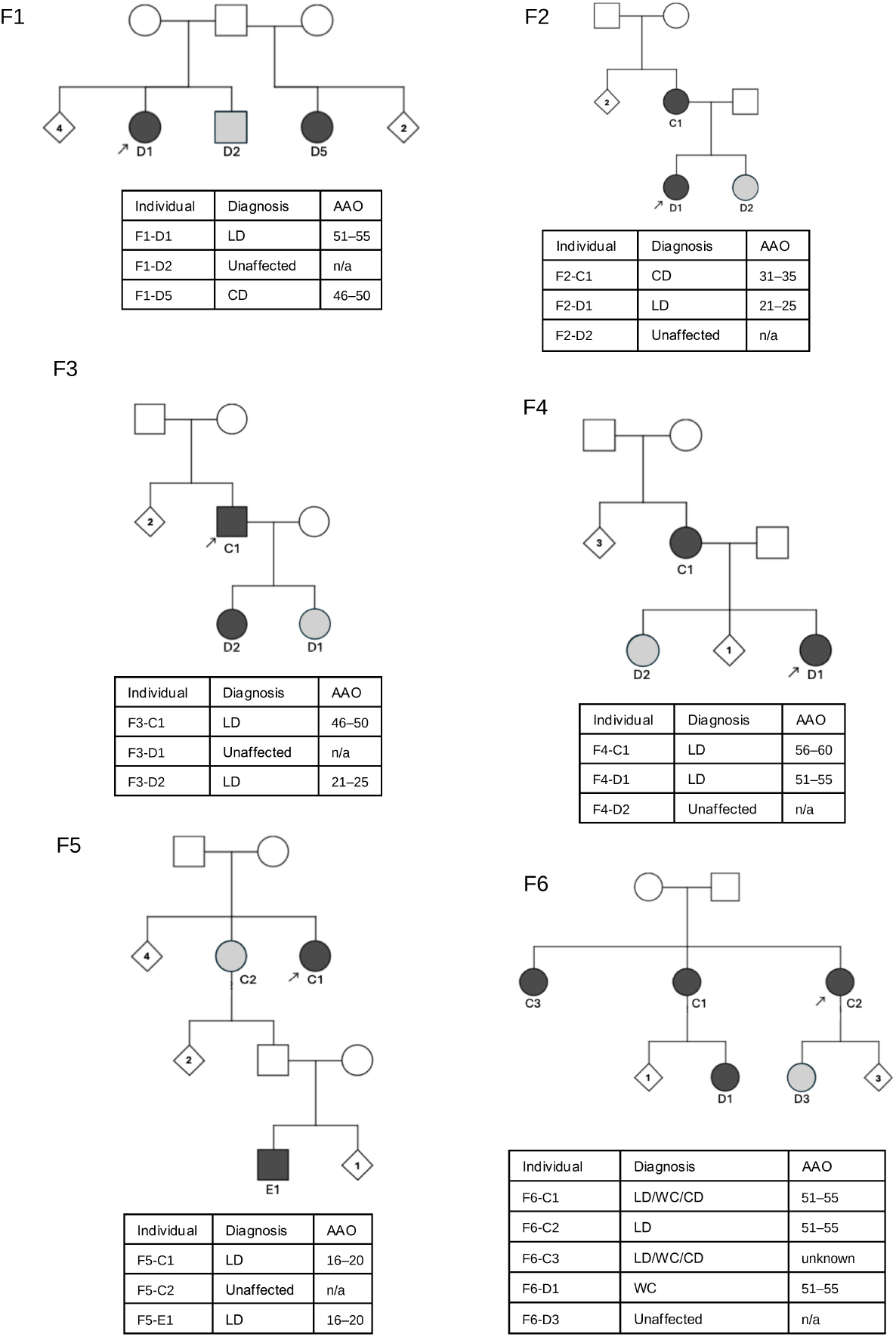
Pedigrees of families in discovery set. DystoniaGeneNet classification of an unaffected relative as a predicted non-manifesting carrier (pNMC) in these families allowed segregation analyses. Affected individuals shaded in black, and pNMCs shaded in grey. Pedigrees have been simplified for privacy of families and uninformative individuals are marked as diamonds. Arrows mark index patient. LD – laryngeal dystonia; CD – cervical dystonia; WC – writer’s cramp; AAO – age-at-onset; NA – not applicable. To protect participant privacy, AAO is shown in 5-year ranges and family identifiers have been recoded.

We further searched for variants in *RAP1GAP, TNN*, and *TMEM190* in replication sets consisting of: (i) the entire WES cohort, consisting of 887 individuals with isolated dystonia, and (ii) a hospital-based biobank (MGBB) where we identified 892 individuals with EHR-defined dystonia. We found 17 additional carriers of rare genetic variation in *RAP1GAP* and *TNN* (**Table 2** [WES], **Supplementary Table 2** [MGBB]); in contrast, no additional families were found to have rare variants in *TMEM190*.

**Table 2.** Variants discovered in *RAP1GAP, TNN, ITSN2*.

| ID | Age/Sex | Diagnosis | AAO | Family history | Variant (HGVS) | Consequence | HGVSp | LOF | AM | CADD | REVEL | gnomAD v4.1 AF | All of Us v9 AF | ClinVar | ACMG |
| --- | --- | --- | --- | --- | --- | --- | --- | --- | --- | --- | --- | --- | --- | --- | --- |
| ***RAP1GAP* (Transcript = ENST00000374765.9, Protein = ENSP00000363897.4)** |  |  |  |  |  |  |  |  |  |  |  |  |  |  |  |
| F1-D1 | 56–60 / F | LD | 51–55 |  |  |  |  |  |  |  |  |  |  |  |  |
| F1-D2 | 61–65 / M | unaffected | n/a | yes | c.1429-1G>A | splice acceptor loss | - | HC | - | 35 | - | - | - | - | LP |
| F1-D5 | 66–70 / F | CD | 46–50 |  |  |  |  |  |  |  |  |  |  |  |  |
| F7-D1 | 76–80 / M | LD | 21–25 |  |  |  |  |  |  |  |  |  |  |  |  |
| F7-D2 | 66–70 / F | unaffected | n/a | yes | c.1801C>T | missense | p.Pro601Ser | - | 0.10 | 24 | 0.43 | $2.6 \times 10^{-5}$ | $9.3 \times 10^{-6}$ | - | VUS |
| F7-E1 | 46–50 / F | LD | 26–30 |  |  |  |  |  |  |  |  |  |  |  |  |
| S1 | 46–50 / F | WC | 41–45 | no | c.41G>A | missense | p.Arg14His | - | 0.84 | 33 | 0.47 | $1.8 \times 10^{-6}$ | $9.3 \times 10^{-6}$ | - | LP |
| S2 | 71–75 / M | CD | 26–30 | no | c.115C>T | nonsense | p.Arg39Ter | HC | - | 48 | - | $5.6 \times 10^{-6}$ | $2.0 \times 10^{-5}$ | - | LP |
| S3 | 66–70 / M | LD | 21–25 | no | c.991C>T | missense | p.Leu331Phe | - | 0.07 | 20 | 0.41 | $1.3 \times 10^{-5}$ | $3.0 \times 10^{-5}$ | VUS | VUS |
| S4 | 46–50 / F | LD | 21–25 | no | c.1270G>A | missense | p.Gly424Ser | - | 0.08 | 17 | 0.61 | $8.6 \times 10^{-6}$ | $1.7 \times 10^{-5}$ | VUS | VUS |
| S5 | >90 / F | CD | 66–70 | no | c.1858A>G | missense | p.Thr620Ala | - | 0.05 | 15 | 0.28 | $6.1 \times 10^{-7}$ | - | - | VUS |
| ***TNN* (Transcript = ENST00000239462.9, Protein = ENSP00000239462.4)** |  |  |  |  |  |  |  |  |  |  |  |  |  |  |  |
| F3-C1 | 61–65 / M | LD | 46–50 |  |  |  |  |  |  |  |  |  |  |  |  |
| F3-D2 | 21–25 / F | LD | 21–25 | yes | c.3647A>G | missense | p.Asp1216Gly | - | 0.98 | 28 | 0.95 | $6.1 \times 10^{-7}$ | $6.5 \times 10^{-6}$ | - | LP |
| F3-D1 | 26–30 / F | unaffected | n/a |  |  |  |  |  |  |  |  |  |  |  |  |
| F8-D1 | 61–65 / F | LD | 61–65 |  |  |  |  |  |  |  |  |  |  |  |  |
| F8-D2 | 61–65 / F | LD | 46–50 | yes | c.3823C>T | missense | p.Arg1275Cys | - | 0.67 | 33 | 0.80 | $6.2 \times 10^{-6}$ | $1.5 \times 10^{-5}$ | - | LP |
| F8-E1 | 41–45 / F | unaffected | n/a |  |  |  |  |  |  |  |  |  |  |  |  |
| S6 | 66–70 / F | LD | 61–65 | no | c.1658C>T | missense | p.Pro553Leu | - | 0.11 | 23 | 0.28 | $1.3 \times 10^{-5}$ | $9.3 \times 10^{-6}$ | - | VUS |
| S7 | 86–90 / F | CD | 36–40 | no | c.1997T>C | missense | p.Val666Ala | - | 0.14 | 24 | 0.42 | $9.2 \times 10^{-6}$ | $6.5 \times 10^{-6}$ | - | VUS |
| S8 | >90 / F | CD | 16–20 | yes | c.2267G>C | missense | p.Gly756Ala | - | 0.16 | 16 | 0.04 | $4.7 \times 10^{-5}$ | $2.5 \times 10^{-5}$ | - | VUS |
| S9 | 66–70 / M | MD | 56–60 | no | c.2570C>T | missense | p.Pro857Leu | - | 0.65 | 24 | 0.62 | $1.9 \times 10^{-5}$ | $3.6 \times 10^{-5}$ | VUS | VUS |
| ***ITSN2* (Transcript = ENST00000355123.9, Protein = ENSP00000347244.4)** |  |  |  |  |  |  |  |  |  |  |  |  |  |  |  |
| S10<sup>1</sup> | 61–65 / F | LD | 31–35 | no | c.3386-2A>T | splice acceptor loss | - | HC | - | 33 | - | $3.1 \times 10^{-6}$ | $3.7 \times 10^{-6}$ | - | LP |
| S11 | 51–55 / F | LD | 31–35 | no | c.4573C>T | nonsense | p.Arg1525Ter | HC | - | 48 | - | $6.1 \times 10^{-6}$ | $3.0 \times 10^{-5}$ | - | LP |
| S12 | 71–75 / F | LD | 51–55 | yes | c.4210C>T | nonsense | p.Arg1404Ter | HC | - | 39 | - | $9.9 \times 10^{-6}$ | $6.5 \times 10^{-6}$ | - | LP |
| S13 | 46–50 / F | LD | 21–25 | no | c.4147G>A | missense | p.Glu1383Lys | - | 0.39 | 26 | 0.227 | $5.0 \times 10^{-5}$ | $2.1 \times 10^{-5}$ | VUS | VUS |
| S14 | 46–50 / F | LD | 36–40 | no | c.4133T>G | missense | p.Leu1378Arg | - | 0.99 | 26 | 0.859 | $2.4 \times 10^{-5}$ | $1.8 \times 10^{-5}$ | - | LP |
| S15 | 76–80 / M | CD | 51–55 | no | c.3139A>G | missense | p.Lys1047Glu | - | 0.32 | 24 | 0.093 | $1.8 \times 10^{-6}$ | - | VUS | VUS |
| S16<sup>1</sup> | 56–60 / M | LD | 51–55 | no | c.2999T>G | missense | p.Leu1000Trp | - | 0.99 | 26 | 0.778 | $1.2 \times 10^{-6}$ | - | - | LP |
| S17 | 31–35 / F | LD | 16–20 | no | c.1616A>G | missense | p.Gln539Arg | - | 0.10 | 24 | 0.096 | $1.9 \times 10^{-6}$ | $5.6 \times 10^{-6}$ | VUS | VUS |
| S18 | 46–50 / F | CD | 26–30 | no | c.1433A>G | missense | p.Gln478Arg | - | 0.17 | 24 | 0.272 | $9.3 \times 10^{-6}$ | $1.0 \times 10^{-5}$ | - | VUS |
| S19 | 51–55 / M | CD | 41–45 | no | c.478C>T | missense | p.Pro160Ser | - | 0.11 | 23 | 0.205 | $3.0 \times 10^{-6}$ | $1.2 \times 10^{-5}$ | VUS | VUS |
| S20 | 66–70 / F | LD | 41–45 | no | c.140A>G | missense | p.Asn47Ser | - | 0.07 | 23 | 0.028 | $1.2 \times 10^{-6}$ | $9.3 \times 10^{-7}$ | - | VUS |

The index genetic variant in *RAP1GAP* was discovered by directly incorporating a pNMC in exome variant filtering in one family with LD. The variant induces a splice acceptor loss in *RAP1GAP*, a gene moderately intolerant to LoF variation (LOEUF = 0.57); further, the variant is absent from large population databases (**Table 2**). To evaluate the predicted LoF associated with this variant, RNA from blood was obtained from a variant carrier, and quantitative PCR was performed. These experiments found loss of *RAP1GAP* expression relative to controls (**Supplementary Fig. 1**). Systematic search for variation in *RAP1GAP* in replication sets identified other genetic variants in isolated dystonia, including a nonsense variant in a patient with sporadic cervical dystonia (CD), missense variants in unrelated individuals with isolated focal dystonia, and a segregating missense variant in a second family with LD (**Table 2**). While some of these variants localize to known functional protein domains, including the catalytic GTPase-activating domain and the regulatory GoLoco motif, others map to the C-terminal phosphoregulatory region^31^ (**Figure 4A**).

**Figure 4.**
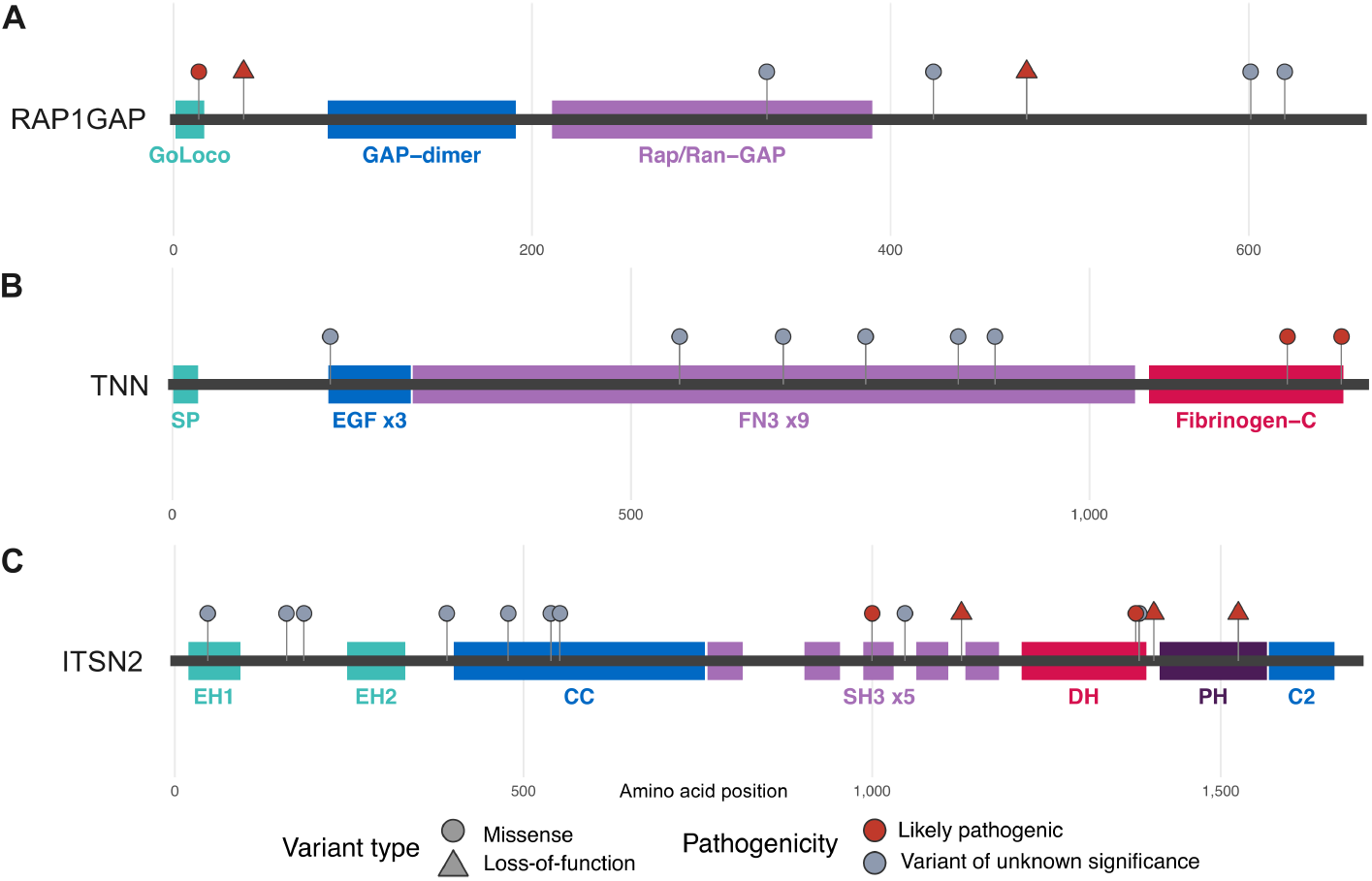
Genetic variants discovered in *RAP1GAP, TNN*, and *ITSN2* in familial and sporadic isolated focal dystonia. Linear protein models of **A**. Rap1GAP **B**. TNN and **C**. ITSN2, showing variants discovered in a dystonia WES cohort and a hospital-based biobank. Coloured boxes indicate protein domains. Variants of unknown significance (VUS) – although these are observed in patients and are rare in population databases, the bioinformatic prediction scores are ambiguous.

The index *TNN* variant was identified in another family with LD, where segregation analysis including a pNMC reduced six candidates to a single likely pathogenic variant. This variant is classified as damaging across all *in silico* prediction methods applied and is ultrarare in the population (**Table 2**). A nearby ultrarare, damaging missense variant in the Fibrinogen-C protein domain was identified in an additional family with LD. Other missense variants in patients with CD were observed in the structural fibronectin type-III repeat domains (**Figure 4B**).

These discovery and replication analyses in families and singletons provide strong genetic evidence that rare LoF and damaging missense variants in *RAP1GAP* and *TNN* contribute to the etiology of isolated focal dystonia.

### Genetic variants in *ITSN2* identified in sporadic isolated dystonia with monogenic features

We further hypothesized that endophenotypes can be used in gene association in sporadic dystonia, by selecting singletons identified by DystoniaGeneNet as exhibiting familial features. We searched for shared genetic risk in this group and identified 11 individuals with a rare genetic variant in *ITSN2*, including three harboring high-confidence LoF variation (**Table 2, Figure 4C**). Further search in the MGBB identified three additional individuals carrying rare missense variants in *ITSN2* (two with CD, one with foot dystonia, **Supplementary Table 2**).

One of the LoF variants identified in *ITSN2* was a splice-altering variant, predicted to result in loss of the canonical splice acceptor for exon 29 and subsequent gain of a new cryptic site in the same exon (SpliceAI^32^ score = 1.0 [acceptor loss], 0.8 [acceptor gain]). We confirmed this effect in blood RNA sequencing data from the variant carrier, where we observed the cryptic splice site to be out-of-frame, introducing a premature stop codon and resulting in loss of 39 amino acids from the C-terminus of *ITSN2* (**Figure 5A**). Exploration of the

**Figure 5.**
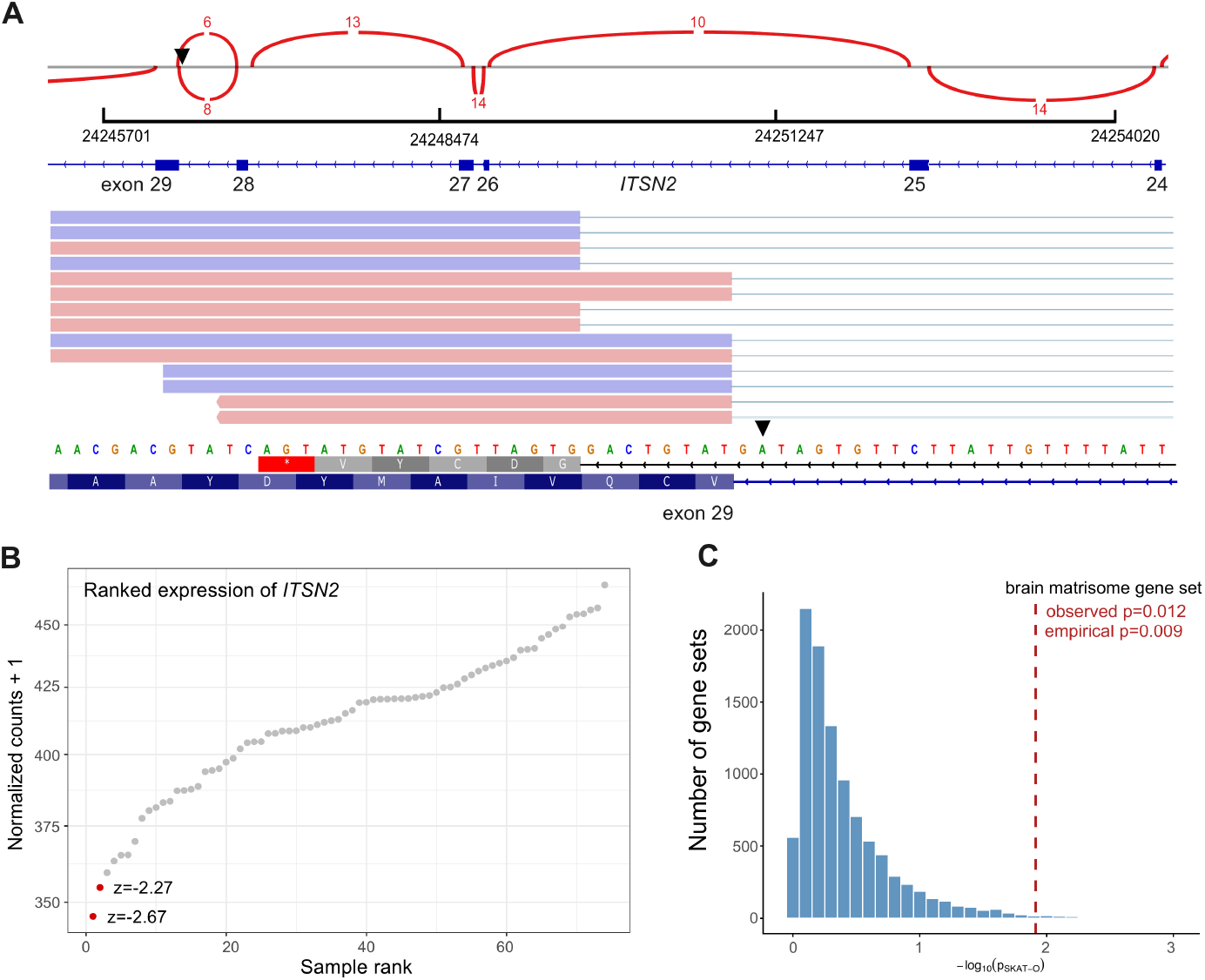
Genetic and transcriptomic evidence of association of *ITSN2* and other matrisome genes with dystonia. **A**. A splice-acceptor variant in *ITSN2* (arrow) results in an *ITSN2* splicing defect as indicated by RNA-Seq splicing analysis and reads. Numbers indicate junction reads and genomic positions. The cryptic splice junction created leads to premature termination of the predicted protein (red box with ^*^) **B**. The splice-acceptor variant and another missense variant in *ITSN2* both result in loss of *ITSN2* expression, and both are outliers across 74 blood RNA-Seq profiles (*P* < 0.05). **C**. Rare nonsynonymous variants collapsed into a gene-set containing brain expressed matrix-related genes (matrisome) are associated with dystonia in a case-control biobank. Competitive test against 10,000 random gene-sets confirmed that this signal is beyond expected by chance alone. *P*-values obtained using Optimized Sequence Kernel Association Tests (SKAT-O).

AlphaGenome Atlas^33^ RNA sequencing dataset revealed a parallel effect in the brain (**Supplementary Fig. 2**). To confirm disruption of gene expression, we performed outlier detection using OUTRIDER^28^ in 74 RNA profiles (**Supplementary Table 3**). This analysis identified reduced expression of *ITSN2* in blood RNA from the splice variant carrier compared with non-carriers of variation in *ITSN2*, and in another individual with sporadic dystonia carrying a missense variant (*z* < -2.0, *P* < 0.05), indicating that both LoF and missense variation induce reduction of *ITSN2* expression in isolated focal dystonia (**Figure 5B**).

In the search for rare variants in *ITSN2* in case-control biobanks, we noted similar rates of missense variation in dystonia and in non-neurologic controls, accompanied by an apparent clustering of likely pathogenic variants towards the C-terminal end of the protein in dystonia (**Figure 4C**). We thus investigated whether specific protein domains are enriched for rare damaging missense variants in cases and performed a within-domain burden analysis using genetic data from 1,343 dystonia cases and 6,840 controls in *All of Us*. For each annotated domain, we compared the number of rare functional missense variants against all other rare variants within the same domain. The Dbl homology (DH) domain of ITSN2 showed significant enrichment of functional missense variants (adjusted *P* = 0.04, **Supplementary Table 4**). No other ITSN2 domain was significantly enriched in dystonia, including the adjacent pleckstrin homology (PH) domain, confirming the signal is specific to the DH domain rather than the broader C-terminus module. No enrichment in dystonia was observed in any domain of Rap1GAP or TNN.

### Association of rare functional variants in perisynaptic matrix genes with dystonia

*RAP1GAP, TNN*, and *ITSN2* do not presently have disease associations in the Online Mendelian Inheritance in Man (OMIM) database (https://omim.org/); however, the proteins they code for all participate in biological processes related to perisynaptic ECM signalling. We thus investigated the overall association of this group of functionally related genes with dystonia via gene burden tests using ancestry-matched genomic data from the MGBB. We selected the core matrisome gene set^34^ as a well-described and validated gene ontology containing genes related to the ECM, additionally refined the set to focus on brain-expressed genes, and to keep this analysis independent of the discovery genes, excluded *RAP1GAP, TNN*, and *ITSN2* from the analysis. While no single matrisome gene was associated with dystonia at exome-wide significance (**Supplementary Table 5**), a pathway-level test detected an association between collapsed rare coding nonsynonymous variants within this gene set and dystonia (*P* = 0.012, *n* = 386 cases and 1,194 controls carrying ≥1 qualifying variant). The robustness of this signal was assessed against 10,000 permutations of random gene sets (of 82 genes each), confirming an association result that was beyond what would be expected by chance (empirical *P* = 0.009, **Figure 5C**).

## Discussion

Gross brain pathologic studies of isolated dystonia do not typically reveal macroscopic abnormalities. Nevertheless, isolated dystonia is considered a brain network disorder due to microstructural, functional, and neurotransmitter alterations in cortical sensorimotor, basal ganglia, thalamic, and cerebellar regions.^35,36^ Neuroimaging studies discriminate carriers of known dystonia-associated genes from controls, and also support that NMCs have subtle brain changes, or endophenotypes, prior to or separate from manifest dystonia.^37–39^ Here, we identified novel genes associated with isolated focal dystonia in independent cohorts and biobanks. This discovery was enabled by DystoniaGeneNet, a deep learning-based framework to automatically identify neural biomarkers of familial dystonia from structural MRIs. Phenoconversion of sporadic cases that were classified as having familial features exemplified the model’s performance in prospectively identifying individuals with increased genetic liability based on neuroimaging features.

Biomarkers included the ITG, a region associated with altered sensorimotor processing characteristic of isolated dystonia.^40,41^ Prior studies report structural differences be-tween putative familial and sporadic forms of LD in the ITG as well as associated white matter pathways in the ILF tract, suggesting that these regions harbour hereditary influences in focal dystonia.^42^ In addition, DystoniaGeneNet identified another key feature in the CC, a structure widely implicated across various forms of focal dystonia and likely responsible for abnormal heteromodal, interhemispheric information transfer.^40,42,43^ Importantly, both the ITG and CC were similarly informative in the DystoniaNet diagnostic platform^20^ and the DystoniaBoTXNet algorithm predicting botulinum toxin treatment efficacy in dystonia,^44^ highlighting the relevance of these regions for dystonia diagnosis, heritability, and therapy.

Leveraging these endophenotypes, we were able to identify three candidate genes and a mechanistic pathway for the etiology of isolated focal dystonia. The genes, *RAP1GAP, TNN*, and *ITSN2*, participate in perisynaptic ECM-synapse signalling. Synapse formation and plasticity are complex processes involving the development of axons and dendrites and their interactions with neural ECM.^45^ Disruption of the ECM around synapses affects brain structural integrity and neural transmission, and leads to neurologic disease.^46^ While the identified genes do not currently have disease associations in humans, their known functions in neurodevelopment and perisynaptic ECM regulation lend biological relevance to our genetic findings. We therefore propose disruption of this pathway as a candidate molecular basis for the network-level structural abnormalities and synaptic plasticity deficits in isolated focal dystonia. A fourth gene nominated by our approach, *TMEM190*, was not found in additional families.

Rap1GAP is enriched in striatal medium spiny neurons and is phosphorylated upon dopamine receptor activation, leading to morphological changes in dendritic spines.^47^ One dystonia-associated variant mapped to the GoLoco/G-protein regulatory motif, the conformation of which is critical for the binding of Gα_o_, the Gα-protein subunit disrupted in *GNAO1*-associated encephalopathy and movement disorder.^48^ TNN (tenascin-N) is a member of the tenascin family of ECM glycoproteins that organize the perisynaptic matrix, and specific protein isoforms direct neurite outgrowth and cell migration in neurons.^49^ Its paralogs tenascin-C and tenascin-R regulate synaptic plasticity.^50^ *ITSN2* codes for a component of the multidomain scaffold protein intersectin.^51^ Related to our observation of an increased burden of dystonia-causing variants in the distal DH domain of *ITSN2*, the long isoform (that exclusively contains this domain) is functionally involved in dendritic spine morphogenesis.^52,53^ In mice, intersectin loss has been shown to result in behavioural abnormalities and defective cortico-striatal neurotransmission,^54^ recapitulating neural transmission abnormalities in dystonia.^55^ Together, these three genes implicate the neural structural matrix (*TNN*) and intracellular GTPase-mediated matrix remodelling (*ITSN2, RAP1GAP*) as connected components of perisynaptic ECM signalling that are plausibly dysregulated in dystonia (**Figure 6**).

**Figure 6.**
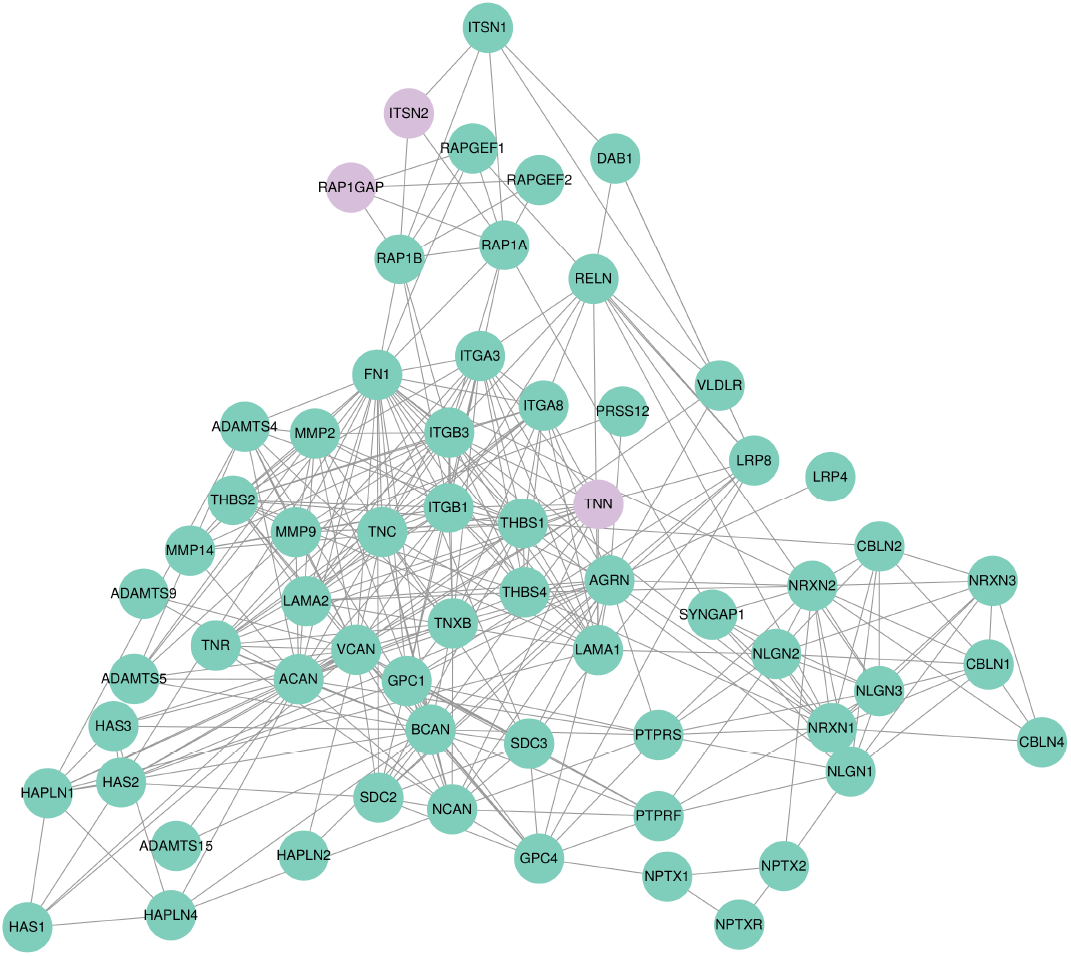
Interaction network of perisynaptic ECM proteins. This STRING database (https://string-db.org/) protein-protein interaction network constructed from proteins that participate in the perisynaptic extracellular matrix (ECM) and its signalling pathways identify dense interactions (edges) between proteins (nodes). Genes identified in dystonia pedigrees and singletons in this study are indicated in purple. Notably, Rap1GAP and ITSN2 cluster in a subnetwork, and have common interaction partners. RELN, also within this network, is a gene associated with myoclonus-dystonia.

We further observed a pathway-level association of rare functional variants in ECM-related genes with isolated dystonia. Genes encoding or regulating brain matrix components have repeatedly surfaced as monogenic causes of dystonia and adjacent movement disorders. These include *COL6A3*, encoding the α3 chain of type VI collagen (early-onset dystonia),^56^ *RELN*, a secreted glycoprotein (myoclonus-dystonia),^57^ and, in mouse models with dystonic phenotypes, Lamb1.^58^ NKX2-1, a transcription factor within the matrisome gene set, regulates neural differentiation in the basal ganglia, and its disruption causes chorea and dystonia.^59^ Finally, in *THAP1*, a frequently observed gene disruption in isolated dystonia pedigrees, our work and that of others implicate dysregulation of lysosomal gene sets and consequent disruption of ECM remodelling and myelin formation in the molecular pathogenesis.^60,61^ Our current findings are consistent with these studies and form a compelling link between genetic variation in perisynaptic ECM-related genes and neural deficits in dystonia.

Our replication sets included samples from biobanks, in which we observed genetic variation in the identified genes in both dystonia patients and in non-neurologic controls. These variants were also found in population databases (gnomAD and *All of Us*), where they were either rare or ultrarare (median allele count of 10; maximum of 82 in gnomAD). This observation is true for other known dystonia-causing variants and reflects the fundamental challenge of dystonia genetics, i.e., that many carriers of pathogenic genetic variants in dystonia-associated genes do not manifest clinically significant phenotypes. For example, even the GAG deletion in *TOR1A*, the most commonly observed mutation in isolated generalized dystonia, is found at a global AF of 5.2×10^-5^, or up to 84 carriers in gnomAD v4.1, reflecting reduced penetrance.^62^ Since our approach to variant filtering integrated NMCs, our discovered variants and genes likely also display reduced penetrance.

Although our sample sizes are suited to family-based segregation analysis and discovery of monogenic disease genes, other models of dystonia genetic etiology should be explored further as cohorts become larger. For example, models in which modifiers of disease expression act in concert with variants in dystonia-associated genes, models incorporating common variation and polygenic risk, and models of oligogenic burden to identify dystonia predisposition genes and their interactions with extragenetic risk should inform the next wave of dystonia genetic association studies.

In summary, we developed DystoniaGeneNet to identify a neural marker of familial dystonia, which we exploited as a neuroimaging endophenotype to advance gene discovery in isolated focal dystonia. The genes we identified converge on the disruption of the perisynaptic ECM as a novel biological mechanism and implicate this pathway as a molecular substrate for the structural and functional deficits in this disorder. Further, our study exemplifies a generalizable strategy for using endophenotypes in genetic discovery, especially in rare neurogenetic diseases with incomplete penetrance and small family sizes.

## Data Availability

Exome data are available via the dbGaP repository (Accession Number: phs004887.v1). Scripts related to WES analysis are available on GitHub (https://github.com/dystonia-omics-lab/dystonia-families).

## Acknowledgements

We thank all participants and their family members. Biospecimens and associated data used in this study were obtained from the Mass General Brigham Biobank (MGBB), under IRB-approved protocol 2009P002312. We also gratefully acknowledge *All of Us* participants for their contributions, without whom this research would not have been possible. We also thank the US National Institutes of Health’s *All of Us* Research Program, for making available the participant data examined in this study.

## Supplementary Figures

**Supplementary Figure 1.**
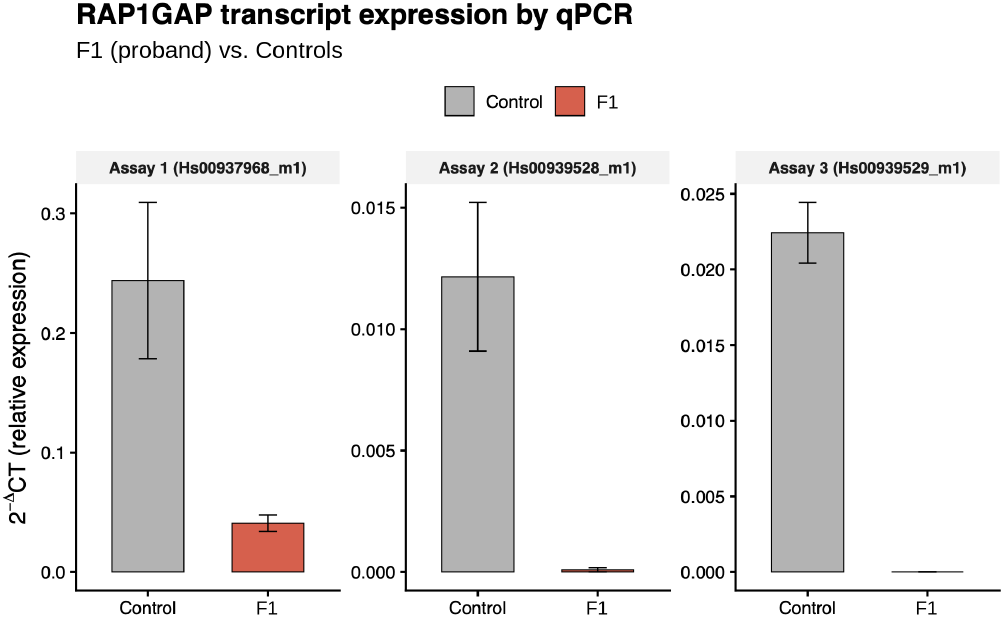
*RAP1GAP* transcript expression by quantitative PCR. Relative expression of *RAP1GAP* was assessed in blood using three Taqman assays and comparison against housekeeping genes. In every assay, transcript expression was almost completely lost in the proband in F1 (carrier of *RAP1GAP* splicing mutation). F1 – bars represent mean of two different runs for each assay. Controls – collapsed from three independent samples. All determinations were done in triplicate.

**Supplementary Figure 2.**
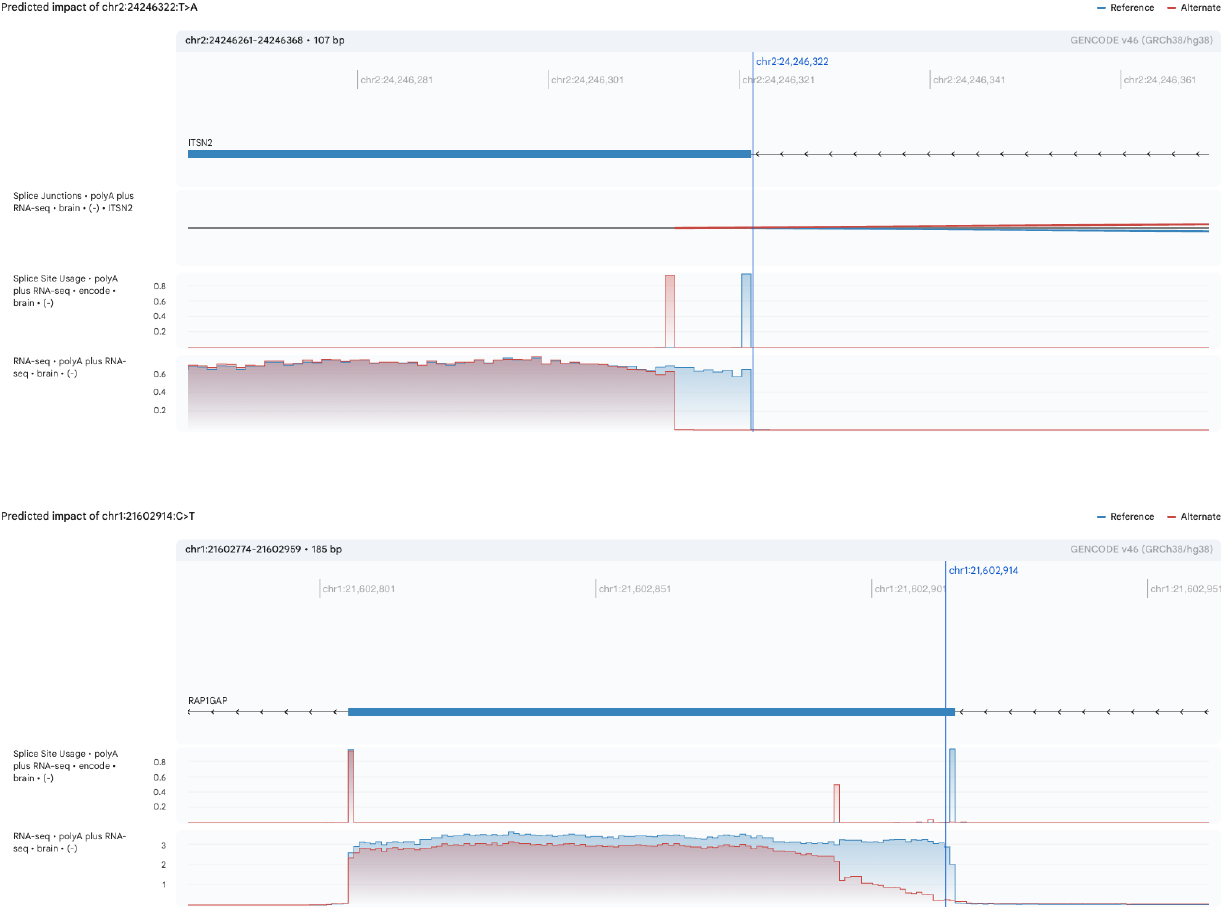
Predicted impact of *ITSN2* and *RAP1GAP* splice variants in brain. AlphaGenome Atlas tracks (https://deepmind.google.com/science/alphagenome/atlas) were explored to determine the impact of splicing variation identified in dystonia individuals in the brain. In both *ITSN2* (top) and *RAP1GAP* (bottom), a cryptic splice site inside the exon is generated because of the splicing variant (red/alternate tracks). In *ITSN2*, the predicted impact in brain parallels the actual observed effect in RNA Sequencing data generated from the patient’s blood (**Figure 5A**).

