## Supplementary Table for "Perisynaptic matrix gene variants underlie DystoniaGeneNet endophenotypes in isolated focal dystonia"

### Contents


| | | |
|---|---|---|
| Supplementary Table 1 | Variant prioritization and segregation. | 2 |
| Supplementary Table 2 | Variants found in Mass General Brigham Biobank. | 7 |
| Supplementary Table 3 | Outlier expression detection of *ITSN2* in blood RNA. | 8 |
| Supplementary Table 4 | Within-domain enrichment tests in *RAP1GAP*, *TNN*, *ITSN2*. | 9 |
| Supplementary Table 5 | Association tests of brain-expressed matrix-related genes (matrisome) with dystonia. | 10 |


### Abbreviations

| | |
|---|---|
| **ACMG** | American College of Medical Genetics variant classification |
| **AF** | Allele Frequency |
| **Bonf** | Bonferroni-corrected p-value |
| **CADD_PHRED** | Phred-scaled score from Combined Annotation Dependent Depletion method |
| **CD** | Cervical dystonia |
| **CI** | Confidence Interval |
| **FDR** | False Discovery Rate |
| **HC** | High Confidence loss of function prediction by LOFTEE |
| **HGVSc** | Variant nomenclature in cDNA based on Human Genome Variant Society |
| **HGVSp** | Variant nomenclature in protein based on Human Genome Variant Society |
| **LOUEF** | Loss of function Observed/Expected Upper bound Fraction |
| **LOFTEE** | Loss of Function Transcript Effect Estimator |
| **LD** | Laryngeal dystonia |
| **LP** | Likely Pathogenic |
| **OR** | Odds Ratio |
| **REVEL** | Rare Exome Variant Ensembl Learner score |
| **SKAT** | Sequence Kernel Association Test |
| **SKATO** | Optimized SKAT (combining SKAT and Burden Test scores) |
| **VUS** | Variant of Unknown Significance |

**Supplementary Table 1.** Variant prioritization and segregation.

| Family | Segregation | Prioritization | Variation | Location | Ref Allele | Alt Allele | Consequence | Gene Symbol | LOEUF | MisZ | HGVSc | HGVSp | LOFTEE Class | Alpha Missense | CADD PHRED | REVEL score | gnomAD4.1 joint AF | AllofUsv9 AF |
|---|---|---|---|---|---|---|---|---|---|---|---|---|---|---|---|---|---|---|
| *Discovery Set* | | | | | | | | | | | | | | | | | | |
| **F1** | **D1 (Aff): 0/1, D5 (Aff): 0/1, D2 (Unaff/pNMC): 0/1** | **Likely Pathogenic** | **chr1:21602914:C:T** | **chr1:21602914** | **C** | **T** | **splice_acceptor_variant** | **RAP1GAP** | **0.593** | **1.368** | **ENST00000374765.9: c.1429-1G>A** | **-** | **HC** | **-** | **35** | **-** | **-** | **-** |
| | | Ambiguous | chr20:885147: GCTGCTGGTC:G | chr20: 885148-885156 | CTGCTGGTC | - | inframe_deletion | ANGPT4 | 1.189 | -0.04688 | ENST00000381922.5: c.757_765del | ENSP00000371347.3: p.Asp253_Gln255del | - | - | 17 | - | $3.1 \times 10^{-6}$ | - |
| | | Likely Benign | chr2:94603893:T:C | chr2:94603893 | T | C | missense_variant | AQP7B | 1.796 | - | ENST00000561882.2: c.719T>C | ENSP00000456868.2: p.Phe241Ser | | 0.309 | 24.8 | - | $2.32 \times 10^{-5}$ | $1.77 \times 10^{-5}$ |
| | | Likely Benign | chr1:21857125:T:C | chr1:21857125 | T | C | missense_variant | HSPG2 | 0.603 | 3.426 | ENST00000374695.8: c.5465A>G | ENSP00000363827.3: p.Asn1822Ser | | 0.2845 | 24.8 | 0.241 | $2.97 \times 10^{-5}$ | $8.4 \times 10^{-6}$ |
| | | Likely Benign | chr5:150387947:G:A | chr5:150387947 | G | A | missense_variant | TCOF1 | 0.288 | 0.9204 | ENST00000643257.2: c.2905G>A | ENSP00000493815.1: p.Ala969Thr | | 0.2001 | 25.8 | 0.388 | $6.51 \times 10^{-5}$ | $3.17 \times 10^{-5}$ |
| | | Likely Benign | chr5:170108361:G:T | chr5:170108361 | G | T | missense_variant | FOXI1 | 1.061 | 0.5745 | ENST00000306268.8: c.887G>T | ENSP00000304286.5: p.Ser296Ile | | 0.1172 | 20.2 | 0.417 | $8.55 \times 10^{-5}$ | $3.64 \times 10^{-5}$ |
| | | Likely Benign | chr5:180630783:C:T | chr5:180630783 | C | T | missense_variant | FLT4 | 0.265 | 3.146 | ENST00000261937.11: c.172G>A | ENSP00000261937.6: p.Glu58Lys | | 0.1723 | 21.7 | 0.09 | $4.86 \times 10^{-5}$ | $6.35 \times 10^{-5}$ |
| | | Likely Benign | chr9:34125190:T:C | chr9:34125190 | T | C | missense_variant | DCAF12 | 0.698 | 1.843 | ENST00000361264.9: c.166A>G | ENSP00000355114.3: p.Arg56Gly | | 0.1179 | 20.1 | 0.202 | $3.72 \times 10^{-6}$ | $9.34 \times 10^{-7}$ |
| | | Likely Benign | chr9:72051821: TAA:T | chr9:72051822-72051823 | AA | - | 3_prime_UTR_variant | C9orf57 | 1.585 | 0.5962 | ENST00000651200.2: c.*473_*474del | - | | - | - | - | - | - |
| | | Likely Benign | chr14:92709830:G:A | chr14:92709830 | G | A | missense_variant | LGMN | 0.852 | 1.273 | ENST00000334869.9: c.862C>T | ENSP00000334052.4: p.Arg288Cys | | 0.0984 | 27.4 | 0.111 | $3.41 \times 10^{-5}$ | $4.95 \times 10^{-5}$ |
| | | Likely Benign | chr17:7442718:A:G | chr17:7442718 | A | G | missense_variant | FGF11 | 1.436 | -0.804 | ENST00000293829.9: c.533A>G | ENSP00000293829.4: p.Glu178Gly | | 0.1144 | 25.9 | 0.425 | $3.1 \times 10^{-5}$ | $1.4 \times 10^{-5}$ |
| **F3** | **C1 (Aff): 0/1, D2 (Aff): 0/1, D1 (Unaff/pNMC): 0/1** | **Likely Pathogenic** | **chr1:175144438:A:G** | **chr1: 175144438** | **A** | **G** | **missense_variant** | **TNN** | **1.089** | **-0.165** | **ENST00000239462.9: c.3647A>G** | **ENSP00000239462.4: p.Asp1216Gly** | | **0.9888** | **28.1** | **0.956** | **$6.2 \times 10^{-7}$** | **$6.53 \times 10^{-6}$** |
| | | Ambiguous | chr8:100630382:T:C | chr8:100630382 | T | C | missense_variant | SNX31 | 1.097 | 0.5771 | ENST00000311812.7: c.266A>G | ENSP00000312368.2: p.Asp89Gly | | 0.3428 | 25.8 | 0.363 | - | - |
| | | Likely Benign | chr1:146126337: GT:G | chr1:146126338 | T | - | frameshift_variant | NBPF10 | 1.966 | -2.481 | ENST00000583866.9: c.1924del | ENSP00000463957.6: p.Thr642LeufsTer8 | HC | - | - | - | $3.83 \times 10^{-6}$ | $7.47 \times 10^{-6}$ |
| | | Likely Benign | chr6:28996539:G:A | chr6:28996539 | G | A | stop_gained | ZNF311 | 0.955 | 1.092 | ENST00000377179.4: c.463C>T | ENSP00000366384.3: p.Gln155Ter | LC | - | 34 | - | - | - |
| | C1: 0/1, D2: 0/1, D1: 0/0 (Sanger) | Likely Pathogenic | chr2:168012632:A:T | chr2:168012632 | A | T | splice_donor_variant | STK39 | 0.381 | 2.076 | ENST00000355999.5: c.1498+2T>A | - | HC | - | 32 | - | - | - |
| | | Ambiguous | chr1:78493095:T:C | chr1:78493095 | T | C | missense_variant | PTGFR | 0.847 | 1.419 | ENST00000370757.8: c.352T>C | ENSP00000359793.3: p.Ser118Pro | | 0.8234 | 26.7 | 0.289 | $2.48 \times 10^{-6}$ | $8.4 \times 10^{-6}$ |
| | | Ambiguous | chr19:40378025:C:T | chr19:40378025 | C | T | missense_variant | PLD3 | 0.727 | 0.8487 | ENST00000409735.9: c.1325C>T | ENSP00000386938.3: p.Ala442Val | | 0.6537 | 28.3 | 0.554 | $1.12 \times 10^{-5}$ | $1.12 \times 10^{-5}$ |
| | | Ambiguous | chr1:78493029:A:C | chr1:78493029 | A | C | missense_variant | PTGFR | 0.847 | 1.419 | ENST00000370757.8: c.286A>C | ENSP00000359793.3: p.Lys96Gln | | 0.2395 | 25.2 | 0.334 | $2.48 \times 10^{-6}$ | $8.4 \times 10^{-6}$ |
| | C1: 0/1, D2: 0/1; not tested in D1 | Likely Benign | chr11:34133141:A:T | chr11:34133141 | A | T | missense_variant,splice_region_variant | NAT10 | 0.935 | 1.472 | ENST00000257829.8: c.1733A>T | ENSP00000257829.3: p.Gln578Leu | | 0.8307 | 31 | 0.525 | $1.43 \times 10^{-5}$ | $1.87 \times 10^{-5}$ |
| | | Likely Benign | chr13:25169665:C:T | chr13:25169665 | C | T | missense_variant | AMER2 | 0.655 | -1.138 | ENST00000515384.2: c.1955G>A | ENSP00000426528.1: p.Arg652Gln | | 0.191 | 28.6 | 0.2 | $7.32 \times 10^{-5}$ | $8.87 \times 10^{-5}$ |
| | | Likely Benign | chr2:235040897:C:A | chr2:235040897 | C | A | missense_variant | SH3BP4 | 0.658 | 1.726 | ENST00000392011.7: c.128C>A | ENSP00000375867.2: p.Pro43His | | 0.2719 | 27.6 | 0.442 | $8.44 \times 10^{-5}$ | $1.78 \times 10^{-4}$ |
| | | Likely Benign | chr6:6145707:C:T | chr6:6145707 | C | T | missense_variant | F13A1 | 0.734 | 1.896 | ENST00000264870.8: c.2111G>A | ENSP00000264870.3: p.Arg704Gln | | 0.1262 | 25.2 | 0.486 | $1.24 \times 10^{-5}$ | $3.64 \times 10^{-5}$ |
| | | Likely Benign | chr11:45910915:C:A | chr11:45910915 | C | A | missense_variant | PEX16 | 0.957 | 0.6338 | ENST00000378750.10: c.935G>T | ENSP00000368024.5: p.Gly312Val | | 0.1646 | 25.1 | 0.356 | - | $1.87 \times 10^{-6}$ |
| | | Likely Benign | chr2:218643172:T:C | chr2:218643172 | T | C | missense_variant | ZNF142 | 0.832 | 2.144 | ENST00000449707.5: c.3344A>G | ENSP00000408643.1: p.Gln1115Arg | | 0.59 | 24.7 | 0.072 | $1.67 \times 10^{-5}$ | $2.24 \times 10^{-5}$ |
| | | Likely Benign | chr3:171167241:C:T | chr3:171167241 | C | T | missense_variant | TNIK | 0.338 | 4.594 | ENST00000436636.7: c.803G>A | ENSP00000399511.2: p.Ser268Asn | | 0.1126 | 24.7 | 0.389 | - | - |
| | | Likely Benign | chr10:119588156: G:C | chr10: 119588156 | G | C | missense_variant | TIAL1 | 0.213 | 4.247 | ENST00000436547.7: c.125C>G | ENSP00000394902.2: p.Thr42Arg | | 0.4536 | 24.4 | 0.266 | - | - |

*continued on next page*

### Supplementary Table 1 (continued)

| Family | Segregation | Prioritization | Variation | Location | Ref Allele | Alt Allele | Consequence | Gene Symbol | LOEUF | MisZ | HGVSc | HGVSp | LOFTEE Class | Alpha Missense | CADD PHRED | REVEL score | gnomAD4.1 joint AF | AllofUsv9 AF |
|---|---|---|---|---|---|---|---|---|---|---|---|---|---|---|---|---|---|---|
| | | Likely Benign | chr2:127505512:T:C | chr2:127505512 | T | C | missense_variant | IWS1 | 0.374 | 1.69 | ENST00000295321.9: c.391A>G | ENSP00000295321.4: p.Ser131Gly | | 0.0892 | 23.4 | 0.111 | $7.87 \times 10^{-5}$ | $5.98 \times 10^{-5}$ |
| | | Likely Benign | chr11:57415044:G:A | chr11:57415044 | G | A | missense_variant | SLC43A3 | 0.804 | 1.316 | ENST00000395124.6: c.832C>T | ENSP00000378556.1: p.Arg278Cys | | 0.1123 | 23.1 | 0.27 | $3.47 \times 10^{-5}$ | $5.04 \times 10^{-5}$ |
| | | Likely Benign | chr6:158663311:C:T | chr6:158663311 | C | T | missense_variant | SYTL3 | 1.342 | -0.7507 | ENST00000611299.5: c.43C>T | ENSP00000483936.1: p.Arg15Cys | | 0.1977 | 22.2 | 0.311 | $5.89 \times 10^{-5}$ | $3.17 \times 10^{-5}$ |
| | | Likely Benign | chr15:54237666:G:C | chr15:54237666 | G | C | missense_variant | UNC13C | 0.857 | -0.7311 | ENST00000260323.16: c.3204G>C | ENSP00000260323.11: p.Arg1068Ser | | 0.575 | 22.2 | 0.602 | $2.17 \times 10^{-5}$ | $7.93 \times 10^{-5}$ |
| | | Likely Benign | chr22:20430717:C:T | chr22:20430717 | C | T | missense_variant | SCARF2 | 0.992 | 0.2851 | ENST00000622235.5: c.1046G>A | ENSP00000477564.2: p.Arg349His | | 0.0953 | 21.4 | - | $3.74 \times 10^{-6}$ | $6.53 \times 10^{-6}$ |
| | | Likely Benign | chr1:109911033:C:T | chr1:109911033 | C | T | missense_variant | CSF1 | 0.338 | 0.6315 | ENST00000329608.11: c.10C>T | ENSP00000327513.6: p.Pro4Ser | | 0.1291 | 21.3 | 0.024 | $3.42 \times 10^{-5}$ | $2.71 \times 10^{-5}$ |
| | | Likely Benign | chr1:45714326:T:C | chr1:45714326 | T | C | missense_variant | IPP | 0.956 | 0.9937 | ENST00000396478.4: c.1450A>G | ENSP00000379739.3: p.Asn484Asp | | 0.1825 | 20.5 | 0.174 | $9.91 \times 10^{-6}$ | $2.61 \times 10^{-5}$ |
| F5 | C1 (Aff): 0/1, E1 (Aff): 0/1, C2 (Obligate/pNMC): 0/1 | Likely Pathogenic | chr17:41524944:G:A | chr17:41524944 | G | A | missense_variant | KRT19 | 0.784 | 0.8049 | ENST00000361566.7: c.559C>T | ENSP00000355124.3: p.Arg187Cys | | 0.2499 | 33 | 0.751 | $4.34 \times 10^{-6}$ | $1.03 \times 10^{-5}$ |
| | | Likely Pathogenic | chr3:47264335:A:C | chr3:47264335 | A | C | missense_variant | KIF9 | 0.873 | 1.436 | ENST00000684063.1: c.932T>G | ENSP00000507186.1: p.Met311Arg | | 0.6922 | 25.6 | 0.511 | $4.96 \times 10^{-6}$ | $8.4 \times 10^{-6}$ |
| | | Likely Benign | chr1:62210848: GAT:G | chr1:62210849-62210850 | AT | - | frameshift_variant | L1TD1 | 1.291 | -1.18 | ENST00000498273.2: c.2078_2079del | ENSP00000419901.1: p.Ile693ArgfsTer5 | LC | - | 20.1 | - | $1.61 \times 10^{-5}$ | $3.27 \times 10^{-5}$ |
| | | Likely Benign | chr1:229654371:G:A | chr1:229654371 | G | A | missense_variant | URB2 | 0.786 | 1.236 | ENST00000258243.7: c.4360G>A | ENSP00000258243.2: p.Val1454Met | | 0.1967 | 21.7 | 0.229 | $2.73 \times 10^{-5}$ | $4.39 \times 10^{-5}$ |
| | | Likely Benign | chr15:42154353:G:C | chr15:42154353 | G | C | missense_variant | PLA2G4F | 1.236 | -0.7119 | ENST00000397272.7: c.290C>G | ENSP00000380442.4: p.Thr97Ser | | 0.1472 | 23.7 | 0.387 | $3.29 \times 10^{-5}$ | $4.48 \times 10^{-5}$ |
| | | Likely Benign | chr19:44977413:G:A | chr19:44977413 | G | A | missense_variant | CLPTM1 | 0.402 | 3.535 | ENST00000337392.10: c.539G>A | ENSP00000336994.4: p.Arg180Gln | | 0.0948 | 23.6 | 0.043 | $1.55 \times 10^{-5}$ | $6.53 \times 10^{-6}$ |
| F2 | C1 (Aff): 0/1, D1 (Aff): 0/1, D2 (Unaff/pNMC): 0/1 | Likely Pathogenic | chr12:12150964:C:T | chr12:12150964 | C | T | missense_variant | LRP6 | 0.27 | 3.989 | ENST00000261349.9: c.2866G>A | ENSP00000261349.4: p.Asp956Asn | | 0.7944 | 33 | 0.424 | $1.24 \times 10^{-6}$ | $1.87 \times 10^{-6}$ |
| | | Likely Pathogenic | chr9:18770619:C:CG | chr9:18770619-18770620 | - | G | frameshift_variant | ADAMTSL1 | 0.744 | -2.496 | ENST00000380548.9: c.2240dup | ENSP00000369921.4: p.Val748CysfsTer5 | HC | - | - | - | - | $9.34 \times 10^{-7}$ |
| | | Likely Pathogenic | chr10:92111116:A:T | chr10:92111116 | A | T | missense_variant | CPEB3 | 0.585 | 1.31 | ENST00000265997.5: c.1532T>A | ENSP00000265997.4: p.Leu511His | | 0.9746 | 27 | 0.415 | - | $1.87 \times 10^{-6}$ |
| | | Likely Pathogenic | chr7:44570849:C:T | chr7:44570849 | C | T | missense_variant | DDX56 | 0.905 | -0.01612 | ENST00000258772.10: c.919G>A | ENSP00000258772.5: p.Gly307Ser | | 0.325 | 28.8 | 0.781 | - | - |
| | | Ambiguous | chr10:129840885: G:T | chr10: 129840885 | G | T | missense_variant | EBF3 | 0.353 | 3.005 | ENST00000440978.2: c.1520C>A | ENSP00000387543.2: p.Pro507His | | 0.743 | 30 | 0.462 | - | - |
| | | Ambiguous | chr19:11174581:G:A | chr19:11174581 | G | A | missense_variant | KANK2 | 0.704 | 0.6219 | ENST00000586659.6: c.1960C>T | ENSP00000465650.1: p.Arg654Trp | | 0.1787 | 29.7 | - | $6.2 \times 10^{-6}$ | $3.73 \times 10^{-6}$ |
| | | Likely Benign | chr6:13791940: TCA:T | chr6:13791941-13791942 | CA | - | frameshift_variant | MCUR1 | 1.334 | -1.704 | ENST00000379170.9: c.960_961del | ENSP00000368468.3: p.Glu321GlyfsTer9 | HC | - | - | - | - | - |
| | | Likely Benign | chr4:158769245:C:G | chr4:158769245 | C | G | missense_variant | FNIP2 | 0.42 | 2.167 | ENST00000264433.11: c.33C>G | ENSP00000264433.6: p.Asn11Lys | | 0.5226 | 25.5 | 0.069 | $3.26 \times 10^{-6}$ | - |
| | | Likely Benign | chr2:182147042: TG:T | chr2:182147043 | G | - | 3_prime_UTR_ variant | PDE1A | 0.857 | 0.6501 | ENST00000409365.6: c.*36del | - | | - | - | - | $7.52 \times 10^{-7}$ | $1.87 \times 10^{-6}$ |
| | | Likely Benign | chr5:71564976: CTG:C | chr5:71564977-71564978 | TG | - | 3_prime_UTR_ variant | BDP1 | 0.683 | 1.345 | ENST00000358731.9: c.*93_*94del | - | | - | - | - | - | $9.34 \times 10^{-6}$ |
| | | Likely Benign | chr12:113321315: G:A | chr12: 113321315 | G | A | missense_variant | SLC8B1 | 1 | 1.232 | ENST00000680972.1: c.190C>T | ENSP00000506377.1: p.Arg64Cys | | 0.1803 | 26.7 | 0.356 | $4.28 \times 10^{-5}$ | $3.08 \times 10^{-5}$ |
| | | Likely Benign | chr17:42787332:C:T | chr17:42787332 | C | T | missense_variant | WNK4 | 0.929 | 1.619 | ENST00000246914.10: c.1531C>T | ENSP00000246914.4: p.Arg511Cys | | 0.8909 | 26.4 | 0.465 | $8.8 \times 10^{-5}$ | $1.22 \times 10^{-4}$ |
| | | Likely Benign | chr5:1079467:C:T | chr5:1079467 | C | T | missense_variant | SLC12A7 | 1.026 | 1.47 | ENST00000264930.10: c.1327G>A | ENSP00000264930.5: p.Asp443Asn | | 0.6988 | 25.8 | 0.753 | $8.12 \times 10^{-5}$ | $9.43 \times 10^{-5}$ |
| | | Likely Benign | chr3:119744137:A:C | chr3:119744137 | A | C | missense_variant | CFAP91 | 1.129 | 0.3801 | ENST00000273390.9: c.1843A>C | ENSP00000273390.5: p.Ser615Arg | | 0.7059 | 24.3 | 0.289 | $2.42 \times 10^{-5}$ | $2.05 \times 10^{-5}$ |
| | | Likely Benign | chr11:119345564: G:C | chr11: 119345564 | G | C | missense_variant | MFRP | 1.307 | -0.9562 | ENST00000619721.6: c.497C>G | ENSP00000481824.1: p.Pro166Arg | | 0.2582 | 23.8 | 0.218 | $4.15 \times 10^{-5}$ | $4.86 \times 10^{-5}$ |

*continued on next page*

### Supplementary Table 1 (continued)

| Family | Segregation | Prioritization | Variation | Location | Ref Allele | Alt Allele | Consequence | Gene Symbol | LOEUF | MisZ | HGVSc | HGVSp | LOFTEE Class | Alpha Missense | CADD PHRED | REVEL score | gnomAD4.1 joint AF | AllofUsv9 AF |
|---|---|---|---|---|---|---|---|---|---|---|---|---|---|---|---|---|---|---|
| | | Likely Benign | chr7:44539795:C:T | chr7:44539795 | C | T | missense_variant | NPC1L1 | 0.955 | 2.2 | ENST00000381160.8: c.602G>A | ENSP00000370552.3: p.Arg201His | | 0.173 | 23.4 | 0.614 | $7.62 \times 10^{-5}$ | $9.89 \times 10^{-5}$ |
| | | Likely Benign | chr9:128987665:A:G | chr9:128987665 | A | G | missense_variant | NUP188 | 0.539 | 2.434 | ENST00000372577.2: c.2341A>G | ENSP00000361658.2: p.Met781Val | | 0.2545 | 23 | 0.276 | $4.34 \times 10^{-6}$ | $1.87 \times 10^{-6}$ |
| | | Likely Benign | chr11:73308889:A:G | chr11:73308889 | A | G | missense_variant | ARHGEF17 | 0.57 | -0.5655 | ENST00000263674.4: c.251A>G | ENSP00000263674.3: p.Gln84Arg | | 0.2603 | 22.5 | 0.105 | $1.62 \times 10^{-5}$ | $2.13 \times 10^{-5}$ |
| | | Likely Benign | chr12:113299892:A:C | chr12:113299892 | A | C | missense_variant | SLC8B1 | 1 | 1.232 | ENST00000680972.1: c.1640T>G | ENSP00000506377.1: p.Leu547Trp | | 0.1598 | 21.2 | 0.382 | $1.05 \times 10^{-5}$ | $1.4 \times 10^{-5}$ |
| | C1: 0/1, D1: 0/1, D2: 0/0 | Likely Pathogenic | chr1:201207261:G:T | chr1:201207261 | G | T | stop_gained | IGFN1 | 0.997 | | ENST00000335211.9: c.2368G>T | ENSP00000334714.4: p.Glu790Ter | HC | - | 32 | - | $1.89 \times 10^{-5}$ | $2.8 \times 10^{-5}$ |
| | | Likely Pathogenic | chr1:203699594:C:G | chr1:203699594 | C | G | missense_variant | ATP2B4 | 0.44 | 0.553 | ENST00000357681.10: c.526C>G | ENSP00000350310.5: p.Gln176Glu | | - | 27.2 | 0.829 | - | - |
| | | Likely Pathogenic | chr14:32546548:C:A | chr14:32546548 | C | A | missense_variant | AKAP6 | 0.367 | 1.019 | ENST00000280979.9: c.1895C>A | ENSP00000280979.4: p.Ala632Glu | | 0.8094 | 25 | 0.363 | - | - |
| | | Likely Pathogenic | chr20:31568082:G:T | chr20:31568082 | G | T | stop_gained | HM13 | 0.564 | | ENST00000398174.9: c.1039G>T | ENSP00000381237.3: p.Glu347Ter | HC | - | 44 | - | - | - |
| | | Ambiguous | chr1:171541743:G:A | chr1:171541743 | G | A | missense_variant | PRRC2C | 0.287 | 0.9458 | ENST00000647382.2: c.4277G>A | ENSP00000495867.2: p.Arg1426His | | - | 26.8 | - | $1.74 \times 10^{-5}$ | $1.21 \times 10^{-5}$ |
| | | Ambiguous | chr1:228175497:C:T | chr1:228175497 | C | T | missense_variant | IBA57 | 1.8 | 1.218 | ENST00000366711.4: c.1055C>T | ENSP00000355672.3: p.Pro352Leu | | 0.3537 | 25.9 | 0.485 | $1.9 \times 10^{-6}$ | $1.96 \times 10^{-5}$ |
| | | Likely Benign | chr1:1437368:G:A | chr1:1437368 | G | A | missense_variant | VWA1 | 1.512 | 1.233 | ENST00000476993.2: c.515G>A | ENSP00000417185.1: p.Arg172Gln | | 0.1174 | 23.7 | 0.497 | $4.09 \times 10^{-5}$ | $4.77 \times 10^{-5}$ |
| | | Likely Benign | chr1:59457019:A:T | chr1:59457019 | A | T | missense_variant | FGGY | 1.2 | 0.999 | ENST00000303721.12: c.613A>T | ENSP00000305922.8: p.Ser205Cys | | 0.1208 | 26.1 | 0.338 | $4.15 \times 10^{-5}$ | $6.44 \times 10^{-5}$ |
| | | Likely Benign | chr1:230322655:GC:G | chr1:230322656 | C | - | 3_prime_UTR_variant | PGBD5 | 0.444 | | ENST00000391860.7: c.*769del | - | | - | - | - | - | - |
| | | Likely Benign | chr2:10788953:G:A | chr2:10788953 | G | A | missense_variant | PDIA6 | 0.619 | 0.8712 | ENST00000272227.8: c.869C>T | ENSP00000272227.4: p.Thr290Met | | 0.1067 | 25 | 0.241 | $3.04 \times 10^{-5}$ | $1.96 \times 10^{-5}$ |
| | | Likely Benign | chr2:53854654:C:A | chr2:53854654 | C | A | missense_variant | GPR75 | 1.395 | 1.055 | ENST00000394705.3: c.103G>T | ENSP00000378195.2: p.Asp35Tyr | | 0.1 | 21.7 | 0.017 | $4.34 \times 10^{-6}$ | $3.73 \times 10^{-6}$ |
| | | Likely Benign | chr3:197514371:T:G | chr3:197514371 | T | G | missense_variant | BDH1 | 1.188 | 0.954 | ENST00000392379.6: c.455A>C | ENSP00000376184.1: p.Glu152Ala | | 0.7304 | 25.9 | 0.638 | $3.47 \times 10^{-5}$ | $4.95 \times 10^{-5}$ |
| | | Likely Benign | chr4:99311521:C:T | chr4:99311521 | C | T | missense_variant,splice_region_variant | ADH1B | 1.013 | 0.8562 | ENST00000305046.13: c.964G>A | ENSP00000306606.8: p.Gly322Ser | | 0.2086 | 26.7 | 0.276 | $1.24 \times 10^{-6}$ | $9.34 \times 10^{-7}$ |
| | | Likely Benign | chr5:172768817:G:A | chr5:172768817 | G | A | missense_variant | DUSP1 | 0.691 | 1.044 | ENST00000239223.4: c.1049C>T | ENSP00000239223.3: p.Thr350Met | | 0.1265 | 25 | 0.137 | $8.89 \times 10^{-6}$ | $1.21 \times 10^{-5}$ |
| | | Likely Benign | chr6:54941344:A:C | chr6:54941344 | A | C | missense_variant | FAM83B | 0.915 | 1.033 | ENST00000306858.8: c.2373A>C | ENSP00000304078.7: p.Lys791Asn | | 0.7655 | 22.2 | 0.082 | $7.25 \times 10^{-5}$ | $7.84 \times 10^{-5}$ |
| | | Likely Benign | chr10:48726016:G:A | chr10:48726016 | G | A | missense_variant | WDFY4 | 0.528 | 1.068 | ENST00000325239.12: c.727G>A | ENSP00000320563.5: p.Val243Met | | 0.291 | 29.6 | 0.274 | $9.67 \times 10^{-6}$ | $1.77 \times 10^{-5}$ |
| | | Likely Benign | chr13:48086407:T:C | chr13:48086407 | T | C | missense_variant | MED4 | 1.033 | 0.9394 | ENST00000258648.7: c.238A>G | ENSP00000258648.2: p.Met80Val | | 0.1933 | 22.7 | 0.242 | $3.72 \times 10^{-6}$ | $4.67 \times 10^{-6}$ |
| | | Likely Benign | chr15:101702261:C:T | chr15:101702261 | C | T | missense_variant | TARS3 | 1.092 | 0.8558 | ENST00000335968.8: c.1199G>A | ENSP00000338093.3: p.Arg400Gln | | 0.2981 | 24.8 | 0.531 | $4.28 \times 10^{-5}$ | $5.69 \times 10^{-5}$ |
| | | Likely Benign | chr17:8370216:C:T | chr17:8370216 | C | T | missense_variant | KRBA2 | 1.68 | 0.8966 | ENST00000396267.3: c.151G>A | ENSP00000379565.3: p.Glu51Lys | | - | 22.1 | 0.068 | - | - |
| | | Likely Benign | chrX:71134801:A:G | chrX:71134801 | A | G | missense_variant | MED12 | - | 0.2545 | ENST00000374080.8: c.4816A>G | ENSP00000363193.3: p.Met1606Val | | 0.0969 | 20.9 | 0.217 | $8.27 \times 10^{-7}$ | - |
| F4 | C1 (Aff): 0/1, D1 (Aff): 0/1, D2 (Unaff/pNMC): 0/1 | Likely Pathogenic | chr6:30656210:A:C | chr6:30656210 | A | C | stop_gained | DHX16 | 0.716 | 3.436 | ENST00000376442.8: c.2486T>G | ENSP00000365625.3: p.Leu829Ter | HC | - | 41 | - | $4.96 \times 10^{-6}$ | $8.41 \times 10^{-6}$ |
| | | Likely Pathogenic | chr6:159249176:T:C | chr6:159249176 | T | C | missense_variant | FNDC1 | 0.676 | -0.5105 | ENST00000297267.14: c.4828T>C | ENSP00000297267.9: p.Phe1610Leu | | 0.9948 | 24.5 | 0.095 | $1.24 \times 10^{-6}$ | $9.34 \times 10^{-6}$ |
| | | Likely Pathogenic | chr17:4596935:G:C | chr17:4596935 | G | C | missense_variant | SMTNL2 | 1.498 | 0.8315 | ENST00000389313.9: c.1065G>C | ENSP00000373964.4: p.Gln355His | | 0.6583 | 23.8 | 0.786 | $5.24 \times 10^{-6}$ | $9.34 \times 10^{-7}$ |
| | | Ambiguous | chr8:141175455:T:C | chr8:141175455 | T | C | missense_variant | DENND3 | 0.805 | 2.394 | ENST00000262585.6: c.2291T>C | ENSP00000262585: p.Ile764Thr | | 0.485 | 26.1 | 0.419 | $3.1 \times 10^{-6}$ | $9.34 \times 10^{-7}$ |
| | | Ambiguous | chr4:8606058:A:G | chr4:8606058 | A | G | missense_variant | CPZ | 1.966 | -6.712 | ENST00000360986.9: c.779A>G | ENSP00000354255.4: p.Tyr260Cys | | 0.3393 | 24.9 | 0.372 | $3.1 \times 10^{-6}$ | $9.34 \times 10^{-7}$ |

*continued on next page*

### **Supplementary Table 1** (continued)

| Family | Segregation | Prioritization | Variation | Location | Ref Allele | Alt Allele | Consequence | Gene Symbol | LOEUF | MisZ | HGVSc | HGVSp | LOFTEE Class | Alpha Missense | CADD PHRED | REVEL score | gnomAD4.1 joint AF | AllofUsv9 AF |
|---|---|---|---|---|---|---|---|---|---|---|---|---|---|---|---|---|---|---|
| | | Ambiguous | chr12:107702961:A:C | chr12:107702961 | A | C | missense_variant | PWP1 | 0.833 | 1.233 | ENST00000412830.8:c.833A>C | ENSP00000387365.3:p.Asp278Ala | | 0.9944 | 28.5 | 0.902 | $2.67 \times 10^{-5}$ | $9.34 \times 10^{-6}$ |
| | | Likely Benign | chr1:201059286:G:A | chr1:201059286 | G | A | missense_variant | CACNA1S | 0.651 | 1.291 | ENST00000362061.4:c.3428C>T | ENSP00000355192.3:p.Ser1143Leu | | 0.3219 | 27.8 | 0.826 | $3.22 \times 10^{-5}$ | $2.71 \times 10^{-5}$ |
| | | Likely Benign | chr4:13579930:G:A | chr4:13579930 | G | A | missense_variant,splice_region_variant | BOD1L1 | 0.412 | 0.9053 | ENST00000040738.10:c.8747C>T | ENSP00000040738.5:p.Thr2916Ile | | 0.21 | 26 | 0.124 | $4.04 \times 10^{-5}$ | $1.77 \times 10^{-5}$ |
| | | Likely Benign | chr5:1216655:G:A | chr5:1216655 | G | A | missense_variant | SLC6A19 | 1.201 | 0.8088 | ENST00000304460.11:c.985G>A | ENSP00000305302.10:p.Ala329Thr | | 0.4426 | 26.7 | 0.862 | $2.48 \times 10^{-5}$ | $2.89 \times 10^{-5}$ |
| | | Likely Benign | chr17:47981470:C:T | chr17:47981470 | C | T | missense_variant | CDK5RAP3 | 0.965 | 1.49 | ENST00000338399.9:c.1489C>T | ENSP00000344683.4:p.Arg497Cys | | 0.7401 | 28.3 | 0.832 | $4.4 \times 10^{-5}$ | $6.81 \times 10^{-5}$ |
| | | Likely Benign | chr22:25722362:G:A | chr22:25722362 | G | A | missense_variant | GRK3 | 0.653 | 2.082 | ENST00000324198.11:c.1979G>A | ENSP00000317578.4:p.Arg660His | | 0.229 | 25.6 | 0.274 | $4.83 \times 10^{-5}$ | $7.09 \times 10^{-5}$ |
| | C1: 0/1, D1: 0/1, D2: 0/0 (Sanger) | Likely Pathogenic | chr8:42290269:G:A | chr8:42290269 | G | A | missense_variant | IKBKB | 0.472 | 0.5549 | ENST00000520810.6:c.314G>A | ENSP00000430684.1:p.Arg105Gln | | 0.9963 | 33 | 0.575 | - | $9.33 \times 10^{-7}$ |
| | | Ambiguous | chr18:63362800:T:C | chr18:63362800 | T | C | missense_variant | KDSR | 0.965 | 0.8421 | ENST00000645214.2:c.177A>G | ENSP00000494352.1:p.Ile59Met | | 0.461 | 21.4 | 0.572 | $2.48 \times 10^{-6}$ | $1.87 \times 10^{-6}$ |
| | | Likely Benign | chr2:53853268:G:C | chr2:53853268 | G | C | missense_variant | GPR75 | 1.395 | 1.055 | ENST00000394705.3:c.1489C>G | ENSP00000378195.2:p.Pro497Ala | | 0.0777 | 21.4 | 0.173 | $1.36 \times 10^{-5}$ | $1.31 \times 10^{-5}$ |
| | | Likely Benign | chr3:8985653:G:T | chr3:8985653 | G | T | missense_variant | SRGAP3 | 0.409 | 0.9596 | ENST00000383836.8:c.3166C>A | ENSP00000373347.3:p.Pro1056Thr | | 0.0826 | 25.4 | 0.075 | $2.26 \times 10^{-5}$ | $1.77 \times 10^{-5}$ |
| | | Likely Benign | chr4:82904759:G:A | chr4:82904759 | G | A | missense_variant | THAP9 | 0.791 | 0.8779 | ENST00000302236.10:c.104G>A | ENSP00000305533.5:p.Arg35His | | 0.7145 | 27 | 0.604 | $8.67 \times 10^{-6}$ | $2.24 \times 10^{-5}$ |
| | C1: 0/1, D1: 0/1; not tested in D2 | Likely Benign | chr5:66144514:G:C | chr5:66144514 | G | C | missense_variant | SREK1 | 0.305 | 1.029 | ENST00000334121.11:c.138G>C | ENSP00000334538.6:p.Glu46Asp | | 0.062 | 20.8 | 0.047 | $1.29 \times 10^{-6}$ | $9.34 \times 10^{-7}$ |
| | | Likely Benign | chr8:27544189:C:A | chr8:27544189 | C | A | missense_variant | EPHX2 | 1.093 | 1.018 | ENST00000521400.6:c.1534C>A | ENSP00000430269.1:p.Pro512Thr | | 0.2149 | 26.2 | 0.542 | $5.45 \times 10^{-5}$ | $2.43 \times 10^{-5}$ |
| F6 | C1 (Aff): 0/1, C2 (Aff): 0/1, C3 (Aff): 0/1, D1 (Aff): 0/1, D3 (Unaff/pNMC): 0/1 | Likely Pathogenic | chr19:55377885:T:C | chr19:55377885 | T | C | missense_variant,splice_region_variant | TMEM190 | 1.413 | -0.2299 | ENST00000291934.4:c.304T>C | ENSP00000291934.3:p.Trp102Arg | | 0.8534 | 25.9 | 0.347 | $2.49 \times 10^{-6}$ | $1.22 \times 10^{-5}$ |
| | | Likely Benign | chr1:15777216:C:T | chr1:15777216 | C | T | missense_variant | FBLIM1 | 1.075 | 0.8017 | ENST00000375766.8:c.937C>T | ENSP00000364921.3:p.Arg313Trp | | 0.143 | 25.5 | 0.269 | $3.91 \times 10^{-5}$ | $3.45 \times 10^{-5}$ |
| | | Likely Benign | chr1:200644441:C:G | chr1:200644441 | C | G | missense_variant | DDX59 | 0.864 | 1.232 | ENST00000331314.11:c.1673G>C | ENSP00000330460.6:p.Trp558Ser | | 0.2877 | 24.6 | 0.122 | - | $1.87 \times 10^{-6}$ |
| | | Likely Benign | chr22:29060630:A:T | chr22:29060630 | A | T | missense_variant | C22orf31 | 1.443 | 0.5143 | ENST00000216071.5:c.217T>A | ENSP00000216071.4:p.Ser73Thr | | 0.1379 | 22.2 | 0.125 | - | - |
| ***Replication*** | | | | | | | | | | | | | | | | | | |
| **F7** | **D1 (Aff), D2 (Oblig), E1 (Aff)** | **Ambiguous** | **chr1:21598478:G:A** | **chr1:21598478** | **G** | **A** | **missense_variant** | **RAP1GAP** | **0.593** | **1.368** | **ENST00000374765.9:c.1801C>T** | **ENSP00000363897.4:p.Pro601Ser** | | **0.1078** | **24.1** | **0.435** | $\mathbf{2.6 \times 10^{-5}}$ | $\mathbf{9.34 \times 10^{-6}}$ |
| | | Ambiguous | chr7:122133533:C:T | chr7:122133533 | C | T | missense_variant | AASS | 0.948 | 1.448 | ENST00000417368.7:c.194G>A | ENSP00000403768.2:p.Arg65Gln | | 0.9654 | 32 | 0.882 | $7.43 \times 10^{-6}$ | $1.4 \times 10^{-5}$ |
| | | Ambiguous | chr7:151042045:G:T | chr7:151042045 | G | T | missense_variant | ABCB8 | 0.894 | 0.9487 | ENST00000297504.10:c.1753G>T | ENSP00000297504.6:p.Ala585Ser | | 0.232 | 23.4 | 0.696 | $4.34 \times 10^{-6}$ | $3.73 \times 10^{-6}$ |
| | | Ambiguous | chr12:18399816:C:T | chr12:18399816 | C | T | missense_variant | PIK3C2G | 1.233 | -1.031 | ENST00000433979.6:c.2161C>T | ENSP00000404845.1:p.Pro721Ser | | 0.238 | 23.8 | 0.516 | $6.25 \times 10^{-7}$ | $1.87 \times 10^{-6}$ |
| | | Likely Benign | chr2:229922518:C:T | chr2:229922518 | C | T | missense_variant | FBXO36 | 1.631 | 0.8333 | ENST00000283946.8:c.5C>T | ENSP00000283946.3:p.Ala2Val | | 0.8012 | 29.1 | 0.159 | $7.44 \times 10^{-6}$ | $1.4 \times 10^{-5}$ |
| | | Likely Benign | chr21:44527320:GC:G | chr21:44527321 | C | - | frameshift_variant | TSPEAR | 1.161 | -0.7384 | ENST00000323084.9:c.1120del | ENSP00000321987.4:p.Ala374ProfsTer38 | HC | - | - | - | $8.05 \times 10^{-6}$ | $1.03 \times 10^{-5}$ |
| | | Likely Benign | chr3:9947708:C:G | chr3:9947708 | C | G | missense_variant | PRRT3 | 1.362 | -2.095 | ENST00000412055.6:c.1465G>C | ENSP00000392511.1:p.Ala489Pro | | 0.7346 | 24.8 | 0.185 | $3.19 \times 10^{-6}$ | $5.6 \times 10^{-6}$ |
| | | Likely Benign | chr11:35432377:C:A | chr11:35432377 | C | A | missense_variant | PAMR1 | 1.274 | 0.432 | ENST00000619888.5:c.2142G>T | ENSP00000483703.1:p.Trp714Cys | | 0.9846 | 32 | - | $2.05 \times 10^{-5}$ | $9.33 \times 10^{-6}$ |
| | | Likely Benign | chr1:32216628:TAG:T | chr1:32216629-32216630 | AG | - | 3_prime_UTR_variant | TMEM234 | 1.63 | 1.021 | ENST00000309777.11:c.*223_*224del | - | | - | - | - | - | - |

*continued on next page*

### **Supplementary Table 1** (continued)

| Family | Segregation | Prioritization | Variation | Location | Ref Allele | Alt Allele | Consequence | Gene Symbol | LOEUF | MisZ | HGVSc | HGVSp | LOFTEE Class | Alpha Missense | CADD PHRED | REVEL score | gnomAD4.1 joint AF | AllofUsv9 AF |
|---|---|---|---|---|---|---|---|---|---|---|---|---|---|---|---|---|---|---|
| | | Likely Benign | chr5:126868495:C:CTAA | chr5:126868495-126868496 | - | TAA | 3_prime_UTR_variant | MARCHF3 | 0.817 | 0.5636 | ENST00000308660.6:c.*2135_*2137dup | - | | - | - | - | - | $1.87 \times 10^{-6}$ |
| | | Likely Benign | chr12:111361942:GAC:G | chr12:111361943-111361944 | AC | - | 3_prime_UTR_variant | PHETA1 | 1.942 | -0.3523 | ENST00000683047.1:c.*734_*735del | - | | - | - | - | - | - |
| | | Likely Benign | chr11:59514973:C:T | chr11:59514973 | C | T | stop_gained | OR4D9 | 1.94 | 0.01604 | ENST00000641962.1:c.61C>T | ENSP00000493010.1:p.Arg21Ter | LC | - | 34 | - | $5.64 \times 10^{-5}$ | $5.41 \times 10^{-5}$ |
| | | Likely Benign | chr14:103534672:C:T | chr14:103534672 | C | T | missense_variant | TRMT61A | 1.183 | 0.9623 | ENST00000389749.9:c.721C>T | ENSP00000374399.4:p.Arg241Cys | | 0.2645 | 31 | 0.456 | $4.66 \times 10^{-5}$ | $8.31 \times 10^{-5}$ |
| | | Likely Benign | chr16:84657855:G:A | chr16:84657855 | G | A | missense_variant | KLHL36 | 0.919 | 0.1894 | ENST00000564996.6:c.1048G>A | ENSP00000456743.1:p.Ala350Thr | | 0.4098 | 26.1 | 0.278 | $2.76 \times 10^{-5}$ | $1.87 \times 10^{-5}$ |
| | | Likely Benign | chr19:3578052:G:T | chr19:3578052 | G | T | missense_variant | HMG20B | 1.087 | -1.385 | ENST00000333651.11:c.880G>T | ENSP00000328269.6:p.Asp294Tyr | | 0.2223 | 25.4 | 0.235 | $6.21 \times 10^{-7}$ | - |
| | | Likely Benign | chr5:142770881:C:G | chr5:142770881 | C | G | missense_variant | ARHGAP26 | 0.493 | 2.886 | ENST00000645722.2:c.120C>G | ENSP00000495131.1:p.Ile40Met | | 0.3577 | 24.8 | 0.259 | $1.24 \times 10^{-6}$ | $9.33 \times 10^{-7}$ |
| | | Likely Benign | chr12:50104635:C:T | chr12:50104635 | C | T | missense_variant | GPD1 | 0.891 | 0.5774 | ENST00000301149.8:c.103C>T | ENSP00000301149.3:p.Arg35Trp | | 0.2454 | 24.7 | 0.265 | $1.74 \times 10^{-5}$ | $3.45 \times 10^{-5}$ |
| | | Likely Benign | chr19:47500690:C:T | chr19:47500690 | C | T | missense_variant | NAPA | 0.538 | 1.26 | ENST00000263354.8:c.238G>A | ENSP00000263354.2:p.Asp80Asn | | 0.5036 | 24.6 | 0.196 | $6.2 \times 10^{-6}$ | $3.73 \times 10^{-6}$ |
| | | Likely Benign | chr3:37324285:T:C | chr3:37324285 | T | C | missense_variant | GOLGA4 | 0.699 | 0.8279 | ENST00000361924.7:c.2399T>C | ENSP00000354486.2:p.Leu800Pro | | 0.4993 | 24 | 0.234 | - | - |
| | | Likely Benign | chr1:17342388:G:T | chr1:17342388 | G | T | missense_variant | PADI4 | 1.012 | 0.5494 | ENST00000375448.4:c.921G>T | ENSP00000364597.4:p.Glu307Asp | | 0.3907 | 23.8 | 0.197 | $6.39 \times 10^{-5}$ | $4.48 \times 10^{-5}$ |
| | | Likely Benign | chr19:45777870:G:A | chr19:45777870 | G | A | missense_variant | DMPK | 0.67 | -0.5587 | ENST00000291270.9:c.679C>T | ENSP00000291270.4:p.Arg227Trp | | 0.132 | 23.8 | 0.234 | $6.66 \times 10^{-5}$ | $7.47 \times 10^{-5}$ |
| | | Likely Benign | chr1:248388384:A:G | chr1:248388384 | A | G | missense_variant | OR2T6 | 1.943 | -0.2916 | ENST00000641644.1:c.776A>G | ENSP00000493366.1:p.Tyr259Cys | | 0.2397 | 23.1 | 0.188 | $6.94 \times 10^{-5}$ | $7.75 \times 10^{-5}$ |
| | | Likely Benign | chr6:17291790:G:A | chr6:17291790 | G | A | missense_variant | RBM24 | 0.612 | 0.1354 | ENST00000379052.10:c.382G>A | ENSP00000368341.5:p.Val128Met | | 0.1587 | 23 | 0.134 | $2.35 \times 10^{-5}$ | $2.33 \times 10^{-5}$ |
| | | Likely Benign | chr1:42191501:G:T | chr1:42191501 | G | T | missense_variant | FOXJ3 | 0.418 | 1.25 | ENST00000361346.6:c.1153C>A | ENSP00000354620.1:p.His385Asn | | 0.0872 | 22.8 | 0.096 | $5.58 \times 10^{-6}$ | $4.67 \times 10^{-6}$ |
| | | Likely Benign | chr17:1726019:C:T | chr17:1726019 | C | T | missense_variant | WDR81 | 0.817 | 0.5577 | ENST00000409644.6:c.1060C>T | ENSP00000386609.1:p.His354Tyr | | 0.1637 | 22.2 | 0.521 | $2.65 \times 10^{-5}$ | $9.33 \times 10^{-6}$ |
| | | Likely Benign | chr10:119827416:C:T | chr10:119827416 | C | T | missense_variant | INPP5F | 0.844 | 1.23 | ENST00000650623.2:c.3035C>T | ENSP00000497527.1:p.Ser1012Leu | | 0.2145 | 21.4 | 0.244 | $5.58 \times 10^{-6}$ | $1.4 \times 10^{-5}$ |
| | | Likely Benign | chr10:6220854:C:T | chr10:6220854 | C | T | missense_variant | PFKFB3 | 0.618 | 1.534 | ENST00000379775.9:c.820C>T | ENSP00000369100.4:p.Arg274Trp | | 0.2168 | 21.3 | 0.637 | $6.95 \times 10^{-5}$ | $1.78 \times 10^{-4}$ |
| | | Likely Benign | chr11:124987226:G:T | chr11:124987226 | G | T | missense_variant | CCDC15 | 1.265 | 0.1512 | ENST00000344762.6:c.1000G>T | ENSP00000341684.5:p.Asp334Tyr | | 0.2241 | 21.3 | 0.211 | $7.09 \times 10^{-6}$ | $8.4 \times 10^{-6}$ |
| | | Likely Benign | chr6:21598150:A:G | chr6:21598150 | A | G | 3_prime_UTR_variant | SOX4 | 0.72 | -1.198 | ENST00000244745.4:c.*2191A>G | - | | - | 20.9 | - | - | $3.73 \times 10^{-6}$ |
| | | Likely Benign | chr17:17143619:C:T | chr17:17143619 | C | T | missense_variant | MPRIP | 0.287 | 3.289 | ENST00000341712.8:c.1099C>T | ENSP00000342379:p.His367Tyr | | 0.074 | 20.7 | 0.066 | - | - |
| | | Likely Benign | chr17:69155890:G:A | chr17:69155890 | G | A | missense_variant | ABCA10 | 1.044 | 0.9632 | ENST00000690296.1:c.3491C>T | ENSP00000509702.1:p.Pro1164Leu | | 0.0891 | 20.7 | 0.364 | $3.66 \times 10^{-5}$ | $4.67 \times 10^{-5}$ |

Variants in bold are reported in Table 2.

**Supplementary Table 2.** Variants found in Mass General Brigham Biobank.

| Gene | Variant HGVS | Consequence | HGVS Protein Change | Protein Domain (UniProt) | LOFTEE | SpliceAI | Alpha Missense | CADD Phred | REVEL | gnomAD4.1 joint AF | AllofUsv9 AF | ClinVar | ACMG | Phenotype |
|---|---|---|---|---|---|---|---|---|---|---|---|---|---|---|
| TNN | c.514C>T | missense | p.Pro172Ser | EGF-like repeat | - | - | 0.1083 | 23.1 | 0.26 | $3.86 \times 10^{-5}$ | $3.55 \times 10^{-5}$ | | VUS | CD, LD |
| TNN | c.2690C>T | missense | p.Thr897Met | Fibronectin type-III 8 | - | - | 0.1291 | 24.2 | 0.561 | $1.67 \times 10^{-5}$ | $4.11 \times 10^{-5}$ | | VUS | LD |
| ITSN2 | c.1655T>C | missense | p.Ile552Thr | Coiled-coil | - | - | 0.1308 | 18.56 | 0.034 | - | $9.34 \times 10^{-7}$ | | VUS | CD |
| ITSN2 | c.1168_ 1170dup | inframe insertion | p.Gln390dup | Coiled-coil | - | - | - | 15.56 | - | - | $1.03 \times 10^{-5}$ | | VUS | CD |
| ITSN2 | c.554C>T | missense | p.Ser185Leu | | - | - | 0.1 | 29.7 | 0.116 | $8.61 \times 10^{-5}$ | $2.08 \times 10^{-4}$ | VUS/Unspecified condition | VUS | foot dystonia |

**Supplementary Table 3.** Outlier expression detection of *ITSN2* in blood RNA.

| ID* | Phenotype | p-value | z-score | log2-foldchange |
|---|---|---|---|---|
| S10 | Dystonia | 0.009056 | -2.67 | -0.25 |
| S16 | Dystonia | 0.02539 | -2.27 | -0.22 |
| | Unaffected | 0.08434 | -2.07 | -0.2 |
| | Unaffected | 0.04094 | -1.91 | -0.18 |
| | Dystonia | 0.0678 | -1.83 | -0.17 |
| | Dystonia | 0.05563 | -1.83 | -0.17 |
| | Dystonia | 0.1891 | -1.64 | -0.16 |
| | Dystonia | 0.2655 | -1.32 | -0.13 |
| | Unaffected | 0.2533 | -1.22 | -0.12 |
| | Dystonia | 0.368 | -1.17 | -0.11 |
| | Dystonia | 0.228 | -1.11 | -0.11 |
| | Dystonia | 0.2946 | -1.09 | -0.1 |
| | Unaffected | 0.5346 | -0.94 | -0.09 |
| | Dystonia | 0.3442 | -0.94 | -0.09 |
| | Dystonia | 0.4213 | -0.93 | -0.09 |
| | Dystonia | 0.4676 | -0.89 | -0.08 |
| | Dystonia | 0.4586 | -0.69 | -0.07 |
| | Dystonia | 0.569 | -0.67 | -0.06 |
| | Dystonia | 0.5253 | -0.64 | -0.06 |
| | Unaffected | 0.7325 | -0.55 | -0.05 |
| | Unaffected | 0.5879 | -0.5 | -0.05 |
| | Unaffected | 0.7134 | -0.37 | -0.04 |
| | Dystonia | 0.8324 | -0.29 | -0.03 |
| | Dystonia | 0.8617 | -0.27 | -0.03 |
| | Dystonia | 0.7969 | -0.27 | -0.03 |
| | Unaffected | 0.882 | -0.16 | -0.02 |
| | Unaffected | 0.9353 | -0.16 | -0.02 |
| | Dystonia | 0.9438 | -0.13 | -0.01 |
| | Dystonia | 0.9328 | -0.13 | -0.01 |
| | Unaffected | 0.9478 | -0.12 | -0.01 |
| | Dystonia | 0.9528 | -0.08 | -0.01 |
| | Dystonia | 0.9601 | -0.08 | -0.01 |
| | Dystonia | 0.9924 | -0.04 | 0 |
| | Dystonia | 1 | -0.01 | 0 |
| | Dystonia | 0.9864 | 0.02 | 0 |
| | Dystonia | 0.9777 | 0.04 | 0 |
| | Dystonia | 0.8961 | 0.12 | 0.01 |
| | Dystonia | 0.8711 | 0.16 | 0.02 |
| | Dystonia | 0.7769 | 0.26 | 0.03 |
| | Dystonia | 0.7868 | 0.27 | 0.03 |
| | Dystonia | 0.8185 | 0.3 | 0.03 |
| | Dystonia | 0.7868 | 0.31 | 0.03 |
| | Dystonia | 0.7808 | 0.31 | 0.03 |
| | Unaffected | 0.7851 | 0.32 | 0.03 |
| | Unaffected | 0.7608 | 0.32 | 0.03 |
| | Dystonia | 0.7383 | 0.32 | 0.03 |
| | Dystonia | 0.7203 | 0.34 | 0.03 |
| | Unaffected | 0.7523 | 0.35 | 0.03 |
| | Dystonia | 0.7202 | 0.36 | 0.03 |
| | Dystonia | 0.6327 | 0.4 | 0.04 |
| | Unaffected | 0.7181 | 0.47 | 0.04 |
| | Unaffected | 0.708 | 0.48 | 0.05 |
| | Dystonia | 0.578 | 0.52 | 0.05 |
| | Dystonia | 0.4959 | 0.59 | 0.06 |
| | Dystonia | 0.4614 | 0.64 | 0.06 |
| | Dystonia | 0.6299 | 0.7 | 0.07 |
| | Dystonia | 0.4496 | 0.74 | 0.07 |
| | Dystonia | 0.4948 | 0.77 | 0.07 |
| | Dystonia | 0.4965 | 0.8 | 0.08 |
| | Unaffected | 0.7145 | 0.82 | 0.08 |
| | Unaffected | 0.4337 | 0.88 | 0.08 |
| | Dystonia | 0.3183 | 0.98 | 0.09 |
| | Dystonia | 0.3374 | 1 | 0.09 |
| | Dystonia | 0.2303 | 1.01 | 0.1 |
| | Unaffected | 0.4798 | 1.14 | 0.11 |
| | Dystonia | 0.3109 | 1.2 | 0.11 |
| | Dystonia | 0.1237 | 1.28 | 0.12 |
| | Dystonia | 0.311 | 1.31 | 0.12 |
| | Dystonia | 0.181 | 1.45 | 0.14 |
| | Unaffected | 0.1126 | 1.49 | 0.14 |
| | Dystonia | 0.1692 | 1.49 | 0.14 |
| | Dystonia | 0.2043 | 1.55 | 0.15 |
| | Unaffected | 0.04829 | 1.58 | 0.15 |
| | Dystonia | 0.1147 | 1.89 | 0.18 |

*only those identified in Table 2 have been assigned IDs in this table

**Supplementary Table 4.** Within-domain enrichment tests in *RAP1GAP*, *TNN*, *ITSN2*.

| Gene | UniProt | Domain | Amino Acids Included | n func vars | n other vars | OR | CI low | CI high | p-value | FDR |
|---|---|---|---|---|---|---|---|---|---|---|
| RAP1GAP | P47736 | N_term_GoLoco | 1-180 | 8 | 12 | 2.101 | 0.6175 | 7.15 | 0.2781 | 0.8342 |
| RAP1GAP | P47736 | Rap_GAP | 181-397 | 9 | 22 | 1.947 | 0.4288 | 8.844 | 0.622 | 0.933 |
| RAP1GAP | P47736 | C_terminal | 398-663 | 6 | 43 | 0.4672 | 0.02589 | 8.43 | 1 | 1 |
| TNN | Q9UQP3 | N_assembly | 1-166 | 1 | 22 | 1.955 | 0.07864 | 48.6 | 1 | 1 |
| TNN | Q9UQP3 | EGF_like | 167-263 | 4 | 15 | 1.421 | 0.218 | 9.264 | 1 | 1 |
| TNN | Q9UQP3 | FN3_repeats | 264-1060 | 25 | 107 | 0.7418 | 0.2346 | 2.345 | 0.7835 | 1 |
| TNN | Q9UQP3 | Fibrinogen_C | 1061-1299 | 8 | 32 | 0.2046 | 0.01147 | 3.652 | 0.2068 | 0.8274 |
| ITSN2 | Q9NZM3 | EH_domains | 1-333 | 8 | 26 | 1.577 | 0.3654 | 6.804 | 0.6596 | 1 |
| ITSN2 | Q9NZM3 | Coiled_coil | 334-756 | 15 | 45 | 1.331 | 0.3964 | 4.472 | 0.7321 | 1 |
| ITSN2 | Q9NZM3 | SH3_cluster | 757-1208 | 7 | 32 | 1.605 | 0.4609 | 5.59 | 0.478 | 1 |
| **ITSN2** | **Q9NZM3** | **DH_domain** | **1209-1433** | **14** | **23** | **11.59** | **1.709** | **78.57** | **0.007162** | **0.04297** |
| ITSN2 | Q9NZM3 | PH_domain | 1434-1551 | 6 | 15 | 0.6533 | 0.0944 | 4.521 | 1 | 1 |
| ITSN2 | Q9NZM3 | C2_domain | 1552-1697 | 10 | 19 | 1.059 | 0.1757 | 6.385 | 1 | 1 |

Bold indicates a significant association.

**Supplementary Table 5.** Association tests of brain-expressed matrix-related genes (matrisome) with dystonia.

| Gene | n variants | Beta | Variance | Burden pval | SKAT pval | SKATO pval | Burden FDR | Burden Bonf | SKAT FDR | SKAT Bonf | SKATO FDR | SKATO Bonf |
|---|---|---|---|---|---|---|---|---|---|---|---|---|
| MATN3 | 14 | 80.04 | 1653 | 0.04903 | 0.01535 | 0.02472 | 0.67 | 1 | 0.9369 | 1 | 0.8172 | 1 |
| PXDN | 40 | -226.2 | 8698 | 0.01531 | 0.3912 | 0.02575 | 0.67 | 1 | 0.9369 | 1 | 0.8172 | 1 |
| SRGN | 2 | 86.76 | 1658 | 0.03312 | 0.06926 | 0.03788 | 0.67 | 1 | 0.9369 | 1 | 0.8172 | 1 |
| SPP1 | 9 | 59.42 | 691 | 0.02381 | 0.06335 | 0.03987 | 0.67 | 1 | 0.9369 | 1 | 0.8172 | 1 |
| COL15A1 | 37 | -119.8 | 3186 | 0.0338 | 0.9234 | 0.0664 | 0.67 | 1 | 0.9482 | 1 | 0.8473 | 1 |
| HSPG2 | 165 | -258.1 | 1.695e+04 | 0.04745 | 0.8576 | 0.09538 | 0.67 | 1 | 0.9433 | 1 | 0.8473 | 1 |
| ZP2 | 14 | -59.31 | 1084 | 0.07158 | 0.9492 | 0.1291 | 0.756 | 1 | 0.9492 | 1 | 0.8473 | 1 |
| CRISPLD1 | 17 | -66.22 | 1608 | 0.09867 | 0.8136 | 0.177 | 0.756 | 1 | 0.9433 | 1 | 0.8473 | 1 |
| EMILIN2 | 19 | 54.49 | 1201 | 0.1159 | 0.1663 | 0.1824 | 0.756 | 1 | 0.9369 | 1 | 0.8473 | 1 |
| LAMA3 | 76 | -147 | 8168 | 0.1038 | 0.6574 | 0.1945 | 0.756 | 1 | 0.9369 | 1 | 0.8473 | 1 |
| COL2A1 | 30 | -88.21 | 3205 | 0.1192 | 0.6617 | 0.2153 | 0.756 | 1 | 0.9369 | 1 | 0.8473 | 1 |
| VWA5B1 | 53 | -108.1 | 4935 | 0.1238 | 0.925 | 0.2263 | 0.756 | 1 | 0.9482 | 1 | 0.8473 | 1 |
| COL6A2 | 34 | 62.02 | 3255 | 0.277 | 0.1314 | 0.2364 | 0.782 | 1 | 0.9369 | 1 | 0.8473 | 1 |
| COL18A1 | 34 | -84.47 | 3162 | 0.133 | 0.8628 | 0.2375 | 0.756 | 1 | 0.9433 | 1 | 0.8473 | 1 |
| NELL2 | 22 | -66.76 | 2042 | 0.1396 | 0.6679 | 0.2455 | 0.756 | 1 | 0.9369 | 1 | 0.8473 | 1 |
| COL5A3 | 52 | -90.76 | 4088 | 0.1558 | 0.8085 | 0.2788 | 0.756 | 1 | 0.9433 | 1 | 0.8473 | 1 |
| SBSPON | 16 | -22.13 | 1655 | 0.5864 | 0.1721 | 0.2883 | 0.9351 | 1 | 0.9369 | 1 | 0.8473 | 1 |
| MGP | 5 | -32.25 | 588 | 0.1835 | 0.6516 | 0.2887 | 0.756 | 1 | 0.9369 | 1 | 0.8473 | 1 |
| EMILIN3 | 32 | -83.56 | 3677 | 0.1682 | 0.8478 | 0.293 | 0.756 | 1 | 0.9433 | 1 | 0.8473 | 1 |
| RSPO4 | 12 | 48.16 | 1278 | 0.1779 | 0.3924 | 0.2933 | 0.756 | 1 | 0.9369 | 1 | 0.8473 | 1 |
| VWA7 | 16 | 48.19 | 1280 | 0.1779 | 0.3097 | 0.2999 | 0.756 | 1 | 0.9369 | 1 | 0.8473 | 1 |
| SPON2 | 15 | -46.38 | 1272 | 0.1935 | 0.833 | 0.3231 | 0.756 | 1 | 0.9433 | 1 | 0.8473 | 1 |
| LTBP1 | 43 | 68.25 | 4093 | 0.2861 | 0.2012 | 0.3481 | 0.782 | 1 | 0.9369 | 1 | 0.8473 | 1 |
| FRAS1 | 103 | -126.9 | 9798 | 0.1998 | 0.8589 | 0.3514 | 0.756 | 1 | 0.9433 | 1 | 0.8473 | 1 |
| COL28A1 | 34 | -62.54 | 2459 | 0.2072 | 0.8546 | 0.3543 | 0.756 | 1 | 0.9433 | 1 | 0.8473 | 1 |
| OMD | 10 | 27.91 | 815.5 | 0.3284 | 0.2235 | 0.3598 | 0.804 | 1 | 0.9369 | 1 | 0.8473 | 1 |
| COCH | 25 | 47.2 | 1800 | 0.2659 | 0.2201 | 0.3686 | 0.782 | 1 | 0.9369 | 1 | 0.8473 | 1 |
| FBN1 | 73 | -112.1 | 8075 | 0.2121 | 0.7485 | 0.3694 | 0.756 | 1 | 0.9433 | 1 | 0.8473 | 1 |
| LAMA5 | 146 | 18.92 | 1.482e+04 | 0.8765 | 0.2098 | 0.3695 | 0.9454 | 1 | 0.9369 | 1 | 0.8473 | 1 |
| HAPLN1 | 7 | -24.14 | 441.5 | 0.2506 | 0.8573 | 0.3811 | 0.782 | 1 | 0.9433 | 1 | 0.8473 | 1 |
| VWA1 | 13 | 41.54 | 1406 | 0.2679 | 0.235 | 0.3865 | 0.782 | 1 | 0.9369 | 1 | 0.8473 | 1 |
| NELL1 | 29 | 40.9 | 3020 | 0.4567 | 0.2358 | 0.3962 | 0.8512 | 1 | 0.9369 | 1 | 0.8473 | 1 |
| NID2 | 25 | -54.12 | 2276 | 0.2566 | 0.747 | 0.4232 | 0.782 | 1 | 0.9433 | 1 | 0.8473 | 1 |
| VIT | 30 | -53.04 | 2254 | 0.2639 | 0.8957 | 0.4328 | 0.782 | 1 | 0.9482 | 1 | 0.8473 | 1 |
| COL13A1 | 29 | 37.13 | 2223 | 0.431 | 0.2667 | 0.4358 | 0.8222 | 1 | 0.9369 | 1 | 0.8473 | 1 |
| CILP | 44 | 59.12 | 4242 | 0.364 | 0.2654 | 0.4417 | 0.8222 | 1 | 0.9369 | 1 | 0.8473 | 1 |
| GAS6 | 19 | 10.04 | 1997 | 0.8223 | 0.2749 | 0.4442 | 0.9454 | 1 | 0.9369 | 1 | 0.8473 | 1 |
| LTBP2 | 48 | 40.11 | 3702 | 0.5097 | 0.268 | 0.4453 | 0.9083 | 1 | 0.9369 | 1 | 0.8473 | 1 |
| IGFBP7 | 4 | 2.84 | 375.2 | 0.8834 | 0.3234 | 0.4621 | 0.9454 | 1 | 0.9369 | 1 | 0.8473 | 1 |
| CHADL | 21 | -42.27 | 1848 | 0.3255 | 0.9144 | 0.5049 | 0.804 | 1 | 0.9482 | 1 | 0.8473 | 1 |
| NTN1 | 25 | 59.69 | 3807 | 0.3333 | 0.3425 | 0.5129 | 0.804 | 1 | 0.9369 | 1 | 0.8473 | 1 |
| COL4A2 | 55 | 34.22 | 5130 | 0.6328 | 0.3175 | 0.5152 | 0.9351 | 1 | 0.9369 | 1 | 0.8473 | 1 |
| VWF | 64 | -73.77 | 5518 | 0.3207 | 0.9402 | 0.52 | 0.804 | 1 | 0.9492 | 1 | 0.8473 | 1 |

*continued on next page*

**Supplementary Table 5** (continued)

| Gene | n variants | Beta | Variance | Burden pval | SKAT pval | SKATO pval | Burden FDR | Burden Bonf | SKAT FDR | SKAT Bonf | SKATO FDR | SKATO Bonf |
|---|---|---|---|---|---|---|---|---|---|---|---|---|
| OGN | 7 | 1.259 | 753.2 | 0.9634 | 0.3486 | 0.5223 | 0.9875 | 1 | 0.9369 | 1 | 0.8473 | 1 |
| THBS1 | 33 | -14.24 | 2767 | 0.7866 | 0.3304 | 0.5275 | 0.9454 | 1 | 0.9369 | 1 | 0.8473 | 1 |
| VCAN | 44 | 19.66 | 4404 | 0.767 | 0.33 | 0.5306 | 0.9454 | 1 | 0.9369 | 1 | 0.8473 | 1 |
| MFAP4 | 10 | 17.34 | 1067 | 0.5955 | 0.3668 | 0.5514 | 0.9351 | 1 | 0.9369 | 1 | 0.8473 | 1 |
| PAPLN | 43 | -56.1 | 3601 | 0.3499 | 0.6535 | 0.5546 | 0.8197 | 1 | 0.9369 | 1 | 0.8473 | 1 |
| EDIL3 | 12 | -25.39 | 859.5 | 0.3864 | 0.5464 | 0.5689 | 0.8222 | 1 | 0.9369 | 1 | 0.8473 | 1 |
| ELN | 16 | -33.77 | 1443 | 0.3741 | 0.4991 | 0.5694 | 0.8222 | 1 | 0.9369 | 1 | 0.8473 | 1 |
| SPOCK2 | 19 | 15.11 | 1869 | 0.7267 | 0.3792 | 0.5817 | 0.9454 | 1 | 0.9369 | 1 | 0.8473 | 1 |
| COL11A2 | 48 | 13.08 | 4825 | 0.8506 | 0.389 | 0.6037 | 0.9454 | 1 | 0.9369 | 1 | 0.8473 | 1 |
| EMID1 | 13 | -24.61 | 914.3 | 0.4156 | 0.7256 | 0.6139 | 0.8222 | 1 | 0.9433 | 1 | 0.8473 | 1 |
| EMILIN1 | 8 | -23.9 | 888.5 | 0.4228 | 0.9035 | 0.6167 | 0.8222 | 1 | 0.9482 | 1 | 0.8473 | 1 |
| GLDN | 12 | 7.093 | 1185 | 0.8367 | 0.4262 | 0.6291 | 0.9454 | 1 | 0.9369 | 1 | 0.8473 | 1 |
| BMPER | 20 | -38.95 | 2421 | 0.4286 | 0.7499 | 0.6395 | 0.8222 | 1 | 0.9433 | 1 | 0.8473 | 1 |
| IGFBPL1 | 10 | -6.669 | 960.2 | 0.8296 | 0.4463 | 0.6461 | 0.9454 | 1 | 0.9369 | 1 | 0.8473 | 1 |
| COL6A1 | 46 | -46.29 | 3457 | 0.4312 | 0.618 | 0.6515 | 0.8222 | 1 | 0.9369 | 1 | 0.8473 | 1 |
| IGFBP3 | 6 | -2.168 | 471.7 | 0.9205 | 0.4831 | 0.6619 | 0.9555 | 1 | 0.9369 | 1 | 0.8473 | 1 |
| IGFBP6 | 1 | -4.453 | 112.1 | 0.6741 | 0.6741 | 0.6741 | 0.9351 | 1 | 0.9369 | 1 | 0.8473 | 1 |
| LGI3 | 18 | 24.44 | 2233 | 0.605 | 0.4632 | 0.6753 | 0.9351 | 1 | 0.9369 | 1 | 0.8473 | 1 |
| AGRN | 74 | 70.09 | 1.081e+04 | 0.5003 | 0.571 | 0.6915 | 0.9083 | 1 | 0.9369 | 1 | 0.8473 | 1 |
| SMOC1 | 13 | 6.806 | 769.7 | 0.8062 | 0.5208 | 0.7165 | 0.9454 | 1 | 0.9369 | 1 | 0.8473 | 1 |
| FBLN2 | 43 | -28.11 | 4259 | 0.6667 | 0.4994 | 0.7263 | 0.9351 | 1 | 0.9369 | 1 | 0.8473 | 1 |
| CRELD1 | 7 | -3.891 | 487.9 | 0.8602 | 0.5491 | 0.73 | 0.9454 | 1 | 0.9369 | 1 | 0.8473 | 1 |
| SLIT2 | 44 | -39.17 | 3717 | 0.5206 | 0.5495 | 0.7473 | 0.9083 | 1 | 0.9369 | 1 | 0.8473 | 1 |
| LGI2 | 22 | -13.64 | 2343 | 0.7781 | 0.5412 | 0.7623 | 0.9454 | 1 | 0.9369 | 1 | 0.8473 | 1 |
| THBS4 | 35 | 38.82 | 4017 | 0.5403 | 0.6451 | 0.7642 | 0.9229 | 1 | 0.9369 | 1 | 0.8473 | 1 |
| SPON1 | 30 | -20.43 | 2917 | 0.7052 | 0.5411 | 0.7652 | 0.9376 | 1 | 0.9369 | 1 | 0.8473 | 1 |
| VTN | 6 | 0.7481 | 853 | 0.9796 | 0.5726 | 0.7671 | 0.9879 | 1 | 0.9369 | 1 | 0.8473 | 1 |
| COL4A1 | 42 | -23.26 | 3882 | 0.7089 | 0.5533 | 0.7793 | 0.9376 | 1 | 0.9369 | 1 | 0.8473 | 1 |
| MFAP1 | 4 | -11.5 | 552.5 | 0.6247 | 0.7967 | 0.7823 | 0.9351 | 1 | 0.9433 | 1 | 0.8473 | 1 |
| COLQ | 13 | -13.89 | 693.8 | 0.5979 | 0.6637 | 0.791 | 0.9351 | 1 | 0.9369 | 1 | 0.8473 | 1 |
| TNXB | 88 | -36.6 | 8101 | 0.6842 | 0.5642 | 0.7951 | 0.9351 | 1 | 0.9369 | 1 | 0.8473 | 1 |
| LAMA1 | 63 | -37.09 | 6693 | 0.6503 | 0.5664 | 0.7964 | 0.9351 | 1 | 0.9369 | 1 | 0.8473 | 1 |
| SPARCL1 | 12 | -14.57 | 1130 | 0.6647 | 0.5926 | 0.7984 | 0.9351 | 1 | 0.9369 | 1 | 0.8473 | 1 |
| CRIM1 | 27 | -23.73 | 2591 | 0.6411 | 0.5907 | 0.8077 | 0.9351 | 1 | 0.9369 | 1 | 0.8473 | 1 |
| AEBP1 | 35 | 7.346 | 3585 | 0.9024 | 0.5972 | 0.8155 | 0.9486 | 1 | 0.9369 | 1 | 0.8473 | 1 |
| INTS14 | 9 | 4.026 | 813.6 | 0.8878 | 0.6405 | 0.8163 | 0.9454 | 1 | 0.9369 | 1 | 0.8473 | 1 |
| MEPE | 6 | -0.3237 | 455.7 | 0.9879 | 0.7137 | 0.8531 | 0.9879 | 1 | 0.9433 | 1 | 0.868 | 1 |
| MATN2 | 18 | 11.84 | 2025 | 0.7925 | 0.6592 | 0.8574 | 0.9454 | 1 | 0.9369 | 1 | 0.868 | 1 |
| AMBN | 9 | 8.793 | 1614 | 0.8267 | 0.7922 | 1 | 0.9454 | 1 | 0.9433 | 1 | 1 | 1 |
